# Predicting the need for escalation of care or death from repeated daily clinical observations and laboratory results in patients with SARS-CoV-2 during 2020: a retrospective population-based cohort study from the United Kingdom

**DOI:** 10.1101/2020.12.14.20248181

**Authors:** Colin J Crooks, Joe West, Andrew Fogarty, Joanne R Morling, Matthew J Grainge, Sherif Gonem, Mark Simmonds, Andrea Race, Irene Juurlink, Steve Briggs, Simon Cruikshank, Susan Hammond-Pears, Timothy R Card

**Affiliations:** Nottingham Digestive Diseases Centre, School of Medicine, University of Nottingham, NG7 2UH; Division of Epidemiology and Public Health, School of Medicine, University of Nottingham, NG5 1PB; NIHR Nottingham Biomedical Research Centre (BRC), Nottingham University Hospitals NHS Trust and the University of Nottingham, NG7 2UH; Nottingham University Hospitals NHS Trust, NG7 2UH; Division of Respiratory Medicine, School of Medicine, University of Nottingham, NG5 1PB

## Abstract

**Objectives:** Currently used prognostic tools for patients with SARS-CoV-2 infection are based on clinical and laboratory parameters measured at a single point in time, usually on admission. We aimed to determine how dynamic changes in clinical and laboratory parameters relate to SARS-CoV-2 prognosis.

**Design:** retrospective, observational cohort study using routinely collected clinical data to model the dynamic change in prognosis of SARS-CoV-2.

**Setting:** a single, large hospital in England.

**Participants:** all patients with confirmed SARS-CoV-2 admitted to Nottingham University Hospitals (NUH) NHS Trust, UK from 1st February 2020 until 30th November 2020.

**Main outcome measures:** Intensive Care Unit (ICU) admission, death and discharge from hospital.

**Statistical Methods:** We split patients into 1st (admissions until 30th June) and 2nd (admissions thereafter) waves. We incorporated all clinical observations, blood tests and other covariates from electronic patient records and follow up until death or 30 days from the point of hospital discharge. We modelled daily risk of admission to ICU or death with a time varying Cox proportional hazards model.

**Results:** 2,964 patients with confirmed SARS-CoV-2 were included. Of 1,374 admitted during the 1st wave, 593 were eligible for ICU escalation, and 466 had near complete ascertainment of all covariates at admission. Our validation sample included 1,590 confirmed cases, of whom 958 were eligible for ICU admission. Our model had good discrimination of daily need for ICU admission or death (C statistic = 0.87 (IQR 0.85-0.90)) and predicted this daily prognosis better than previously published scores (NEWS2, ISCARIC 4C). In validation in the 2nd wave the score overestimated escalation (calibration slope 0.55), whilst retaining a linear relationship and good discrimination (C statistic = 0.88 (95% CI 0.81 −0.95)).

**Conclusions:** A bespoke SARS-CoV-2 escalation risk prediction score can predict need for clinical escalation better than a generic early warning score or a single estimation of risk at admission.

**What is already known on this topic:** SARS-CoV-2 is a recently emerged viral infection, which presents typically with flu like symptoms, can have severe sequelae and has caused a pandemic during 2020.

A number of risk factors for poor outcomes including obesity, age and comorbidity have been recognized.

Risk scores have been developed to stratify risk of poor outcome for patients with SARS-CoV-2 at admission, but these do not take account of dynamic changes in severity of disease on a daily basis.

**What this study adds:** We have developed a dynamic risk score to predict escalation to ICU or death within the next 24 hours.

Our score has good discrimination between those who will and not require ICU admission (or die) in both our derivation and validation cohorts.

Our bespoke SARS-CoV-2 escalation risk prediction score can predict need for clinical escalation better than a generic early warning score or a single estimation of risk at admission.

## Introduction

The SARS-CoV-2 pandemic during 2020 has brought health systems to a state of near collapse in some places[1], has probably increased the risk of death from other diseases due to the diversion of resources[2,3], and has necessitated changes in regulations and behaviour across societies in many parts of the world. However, whilst the pandemic is undergoing further waves of infection the focus again must be on caring for the severely ill with the limited resources available while maintaining care for patients with other problems.

During the first wave of the pandemic massive quantities of data were collected and a number of scores for prioritisation and prognostication were created and assessed[4–6]. Ideal scoring systems would allow safe and early discharge of those that would not be likely to require ongoing hospital care, while at the same time enabling prompt escalation of care for those with deteriorating disease. Although focused primarily on direct patient care, such a system would ensure that resource utilisation was allocated appropriately based on care requirements. Some scores studied like ISARIC 4C[5] are bespoke for SARS-CoV-2 and likely therefore to perform better in this specific condition, but in general these are based on analysis at a single time point (admission), whereas clinical decisions regarding escalation of care must be made by clinicians throughout the disease course. Others are aimed at more dynamic use through the disease course, but are not disease specific (such as NEWS2[7]). A number of these perform reasonably, for example the area under the receiver operating characteristic curve is 0.77 (0.76-0.77) for the validation cohort of ISARIC 4C[5], and for NEWS-2 the AUC varied between hospitals from 0.623 to 0.815[8]. A simple score which is intended to be both dynamic and optimised for SARS-CoV-2 might however be expected to perform better and with levels of disease rising again such a scoring system would be of great value.

In Nottingham our electronic records provide for the collection of contemporaneous, clinical observations and blood tests throughout admission and by linking these to data at presentation we are able to examine retrospectively the performance of scores across the period of an admission. We have therefore set out to develop a truly dynamic and SARS-CoV-2 specific score, to internally validate the score, and to compare and contrast it’s performance to the NEWS2 and ISARIC 4C scores.

## Methods

We carried out and reported this study, in accordance with the TRIPOD guidelines for multivariable prediction of individual prognosis or diagnosis studies[9].

### Study design, setting and populations

This retrospective, observational cohort study was conducted at Nottingham University Hospitals (NUH) NHS Trust, UK. NUH has a catchment population of approximately 2.5 million with 1,500 inpatient hospital beds. Using the electronic health record systems within NUH, all patients suspected of having SARS-CoV-2 who attended or were already inpatients from 21 February 2020 (the date of disease onset of the first known case) until 30 June 2020 inclusive were identified for a derivation cohort. Patients admitted after this date until 30th November 2020 formed a validation cohort. From this cohort, we identified all patients who were considered to have definite SARS-CoV-2 either via a positive result on polymerase chain reaction (PCR) testing of a nasopharyngeal sample or a recorded clinical diagnosis of SARS-CoV-2. We had complete follow up for discharge, subsequent admissions to NUH, and death both in and outside of hospital (via the NHS Patient Demographics Service (PDS) until 30th November 2020.

### Data collection

Data were extracted from the available electronic records (including System C’s Medway Electronic Patient Record system, daily local Intensive Care National Audit & Research Centre returns and NerveCentre http://nervecentresoftware.com/) with the use of an enterprise data warehouse. Information extracted comprised of demographic information, ethnicity, comorbidities, ceiling of care decisions, type of ward each patient was on and for how long (i.e. standard inpatient ward, intensive care unit), oxygen delivery systems and flow, invasive mechanical ventilation, length of stay, readmission, alcohol risk assessment, smoking and/or vaping status, body mass index (BMI) / nutritional assessment. All laboratory tests, results (microbiology and blood sciences) and clinical observations (respiratory rate, blood pressure, pulse rate, oxygen saturation, temperature) carried out from a hospital admission during which SARS-CoV-2 was confirmed, or from date of suspicion of SARS-CoV-2 if preceding the date of admission from an ED attendance; until admission to ICU, discharge from hospital or in-hospital death were extracted for use in the modelling.

### Statistical Analysis

#### Outcome

For those patients eligible for escalation of respiratory support the combined outcome of either of first ICU admission or death was defined from the hospital and NHS PDS for all patients. We assessed whether the outcome occurred on the day of admission and on each subsequent day until 60 days post their earliest suspected date of SARS-CoV-2. Patients were modelled as at risk until 24 hours prior to an outcome. This was to train the model covariates to predict next day events. For those patients ineligible for escalation death alone was defined in similar fashion.

#### Baseline covariates

Ethnic group was categorised as Black/Mixed, Asian, White and other/not stated. Alcohol risk assessment was a binary covariate from admission screening as at risk or not, smoking as a binary variable of a current smoking and/or vaping status, BMI was derived from the nutritional assessment and categorised as BMI <20, 20-30 or >30 kg/m2. Age was categorised as a linear variable, a quadratic transformation and 20 year categories (20-39, 40-59, 60-79, >79 years), and likelihood ratio tests used to select the best fit. The presence of co-morbidity was categorised by the recording of any co-morbidity in the Charlson index[10].

#### Time varying covariates

The daily mean of each blood test was calculated along with the daily change (difference between first and last measurement within a day), and the lagged change in the mean value from the previous day. Last observed measurements were carried forward for calculating the daily summary measures, and patient days prior to any measurements were not included. For multiple observations, the worst daily value was recorded, along with the daily change (difference between first and last measurement within a day), and the lagged change in the worst value from the previous day. For very positively skewed variables a log transformation was applied, and all covariates were centred on their mean for the analysis.

#### Model selection

The de-identified data were analysed using a time varying Cox proportional hazards model. The model was selected using both forward and backward steps using Akaike information criterion (AIC) as a measure of goodness of fit, in addition to a manual assessment of covariates to remove implausible associations from correlated covariates. The process was further assessed with bootstrapping 100 times to show the uncertainty in parameter selection and identify the parameters consistently selected.

#### Sample size

Pragmatically we intended to use all available patients in the cohort to derive and validate the model. However, prior to the study we used the r package pmsampsize to calculate the minimum sample size required to develop a multivariable prediction model for a binary outcome using 10 candidate predictor parameters. Based on previous evidence, the outcome prevalence was anticipated to be 0.12-0.27 and a lower bound for the new model’s acceptable R-squared value as 0.15. This estimated a sample size of around 500 patients.

#### Internal validation and comparison to NEWS2 and ISARIC4C

The performance of the model was assessed using the C statistic fitted with leave one out cross validation, and compared across different time points in the follow up. Optimism introduced through the selection of parameters was adjusted for using the uniform shrinkage factor calculated from bootstrapping the model selection process. We then fitted the derived score from the 1st wave cohort in the 2nd wave cohort and assessed its calibration and performance. Finally, we compared our score with the performance of NEWS2 and ISARIC 4C by implementing these scores using the published methods, and computing their performance characteristics in terms of discrimination and calibration. A sensitivity analysis was undertaken validating with only those patients with confirmatory PCR for SARS-CoV-2.

All analyses were performed using version 4.0.3 of the R programming language (R project for Statistical Computing; R Foundation). Approval for this work was granted via an NUH Clinical Effectiveness Team audit (reference: 20-153C), the NUH Caldicott Guardian, Data Protection Impact Assessment (reference: 436) and as a research study (ethics approval) via the NHS Health Research Authority (HRA) Integrated Research Application System (IRAS) (reference: 282490). The HRA confirmed that individual patient consent was not required for this work.

### Patient and public involvement

The protocol was discussed by our NIHR Biomedical Research Centre Patient Advisory Group that was convened for SARS-CoV-2 projects and the feedback was incorporated.

### Funding

This work was funded by Nottingham University Hospitals NHS Trust and the University of Nottingham. Nottingham University Hospitals NHS Trust also sponsored the study. Neither organisation beyond their employees who are the authors had any role in the design, analysis, interpretation, writing up or submission of this work. All data was collected originally during routine clinical care in Nottingham University Hospitals NHS Trust.

The guarantors affirm that the manuscript is an honest, accurate, and transparent account of the study being reported; that no important aspects of the study have been omitted; and that any discrepancies from the study as originally planned and registered (clinicaltrials.gov <u>NCT04473105</u>) have been explained.

### Data sharing

The individual data used in this study, under the information governance and HRA IRAS approvals, are unable to be shared beyond NUH Hospitals NHS Trust.

## Results

The complete daily status of patients from the combined derivation and validation cohort is shown in Figure 1 by calendar date and their demographic, baseline characteristics and mortality outcomes are shown in table 1. Overall, 2,964 patients were admitted and the key differences apparent between the 1^st^ and 2^nd^ wave cohorts were that in the 2^nd^ wave the median age was slightly lower (1^st^ wave: 76 versus 2^nd^ wave: 73) and 30-day mortality was substantially lower (1^st^ wave: 27% versus 2^nd^ wave: 20%).

**Table 1.**
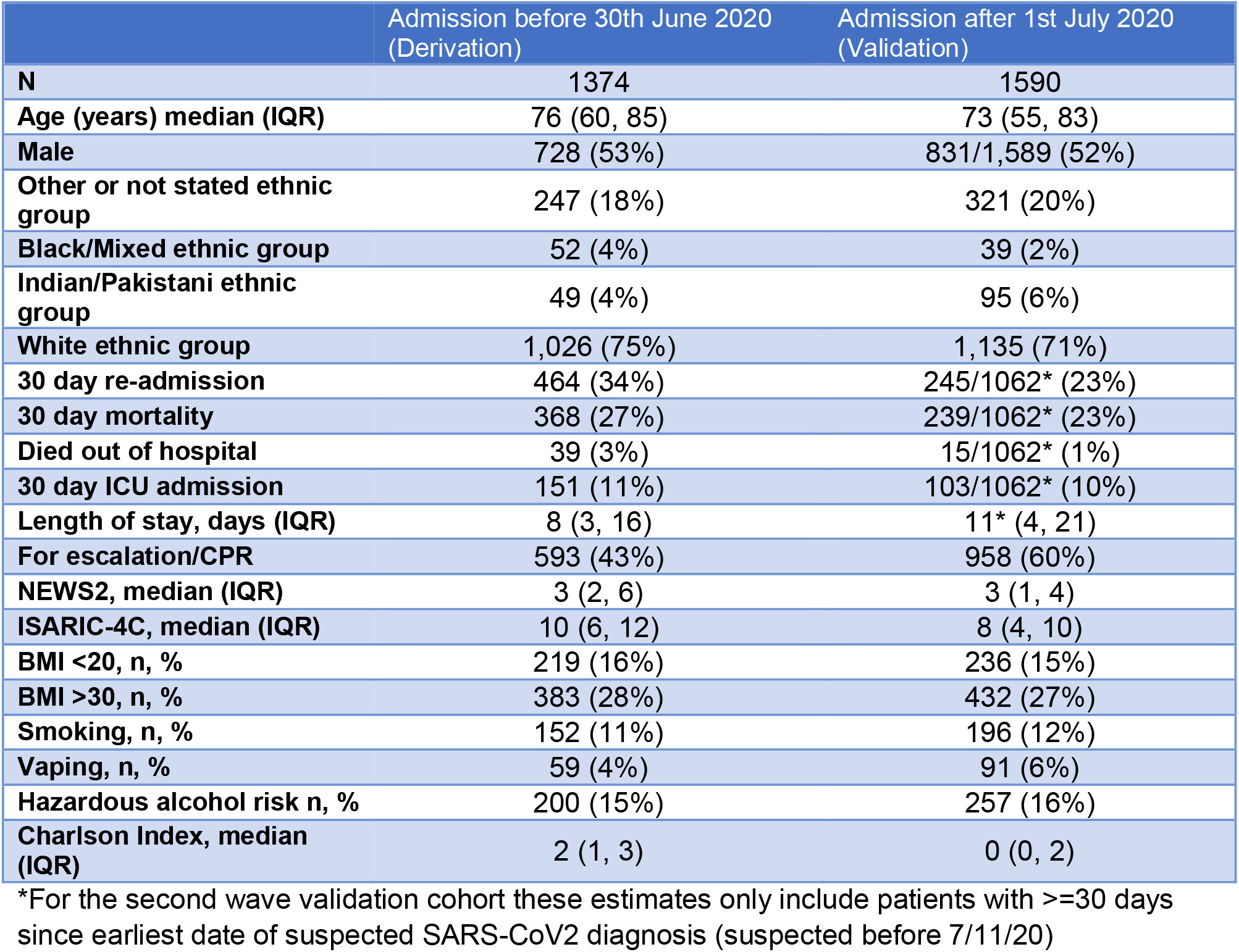
Sociodemographic and other characteristics on admission of the derivation cohort who were admitted to hospital with confirmed SARS-COV-2 diagnosis 21 February 2020 until 30 June 2020

**Figure 1.**
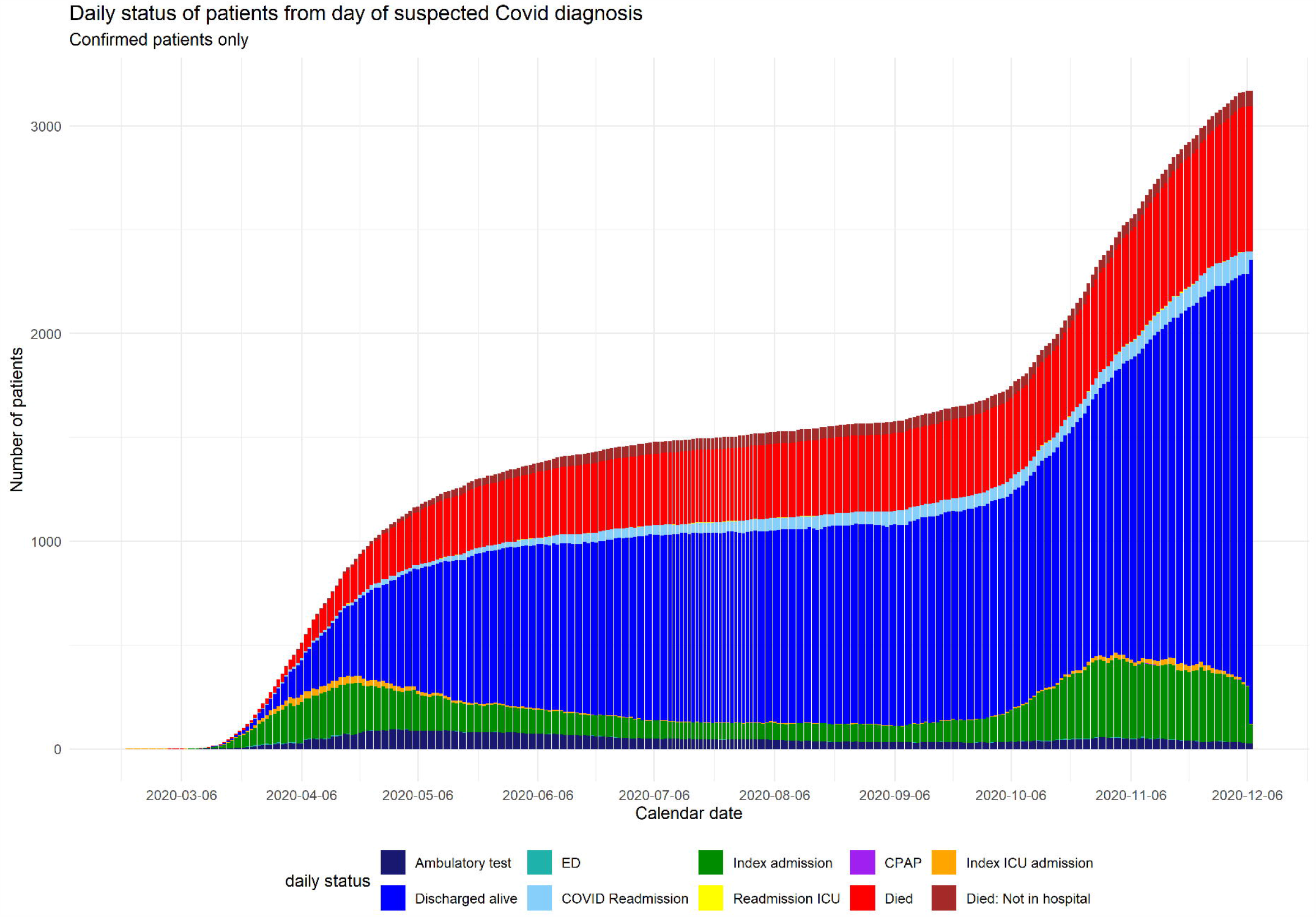
Overall cohort by calendar date.

### First wave derivation cohort

From 21 February 2020 until 30 June 2020 5,879 patients who were initially assessed as suspected SARS-CoV-2 attended Nottingham University Hospitals. During the follow up period 1,449 patients were eventually confirmed clinically as a SARS-CoV-2 diagnosis. The daily status of these patients is shown in figure S1 by day of disease course (measured from the day on which the suspicion of having SARS-CoV-2 was first recorded).

Of those patients with clinical or PCR confirmed SARS-CoV-2 diagnosis 1,374 were admitted of whom 986 (72%) had a confirmatory PCR (supplementary table 1). 593 were eligible for escalation of respiratory support during their initial inpatient period (as determined by the teams caring for them) and 152 were escalated or died within the 30 day follow up period. 582 patients were suspected of having SARS-CoV-2 before any escalation occurred, and 527 during the initial 48 hours (i.e. suspected on admission), with blood tests and observations as shown in the flow charts in figure S2 in supplementary figures.

### Second wave validation cohort

From 1st July 2020 until 30th November 2020 5,674 patients who were initially assessed as suspected SARS-CoV-2 attended Nottingham University Hospitals. During the follow up period 1,590 patients were eventually confirmed clinically as a SARS-CoV-2 diagnosis (882 with PCR positive tests to date) of whom 958 were admitted eligible for escalation, and 632 were admitted ineligible for escalation.

### Patients eligible for escalation in 1^st^ wave: Predicting daily risk of next day ICU admission or death

Table 2 shows initial measurements for patients on admission by final outcome, and figure S3 shows how selected observation and blood results further varied over time. There were initial higher neutrophil counts and oxygen requirements in patients who required escalation or died within 30 days of earliest suspected date of SARS-CoV-2, which gradually reduced in patients who survived as fewer patients remained at risk. Modelling daily summary measures of full blood count, urea and electrolytes, and observations as described in the methods, a model with a quadratic term for age was a statistically better fit (likelihood ratio test p = 0.01 compared to model with only age as a linear term), but the categorisation of age did not improve the model significantly (likelihood ratio test p = 0.5). Blood cell counts were transformed to the log scale due to positive skew. The daily risk of escalation or death was then predicted in a time varying Cox proportional hazards model with the last observed value carried forward. The final selected model is shown in the second column of table 3, and the adjustment for optimism using the bootstrapped uniform shrinkage factor was estimated at 0.73 (IQR 0.64-0.80). The proportion of bootstrapped samples within which a covariate was selected is shown in the third column in table 3.

**Table 2.** Initial blood test results and observations on first admission of confirmed SARS-COV-2 patients by eligibility for escalation of care and worst outcome 21 February 2020 until 30 June 2020 (derivation cohort)

|  | Not for escalation |  | Eligible for escalation |  |  |
| --- | --- | --- | --- | --- | --- |
|  | Survived | Died | Not escalated | ICU admission | Died |
| **Number of patients suspected within 48 hours of admission** | 324 | 450 | 395 | 112 | 75 |
| **Haemoglobin, median (IQR)** | 122 (110, 132) | 116 (100, 133) | 129 (116, 142) | 121 (107, 131) | 121 (104, 133) |
| **Not initially measured** | 45 (14%) | 66 (15%) | 52 (13%) | 3 (3%) | 6 (8%) |
| **Platelets, median (IQR)** | 217 (167, 290) | 219 (156, 280) | 219 (176, 290) | 246 (197, 323) | 236 (156, 332) |
| **Not initially measured** | 54 (17%) | 93 (21%) | 71 (18%) | 8 (7%) | 10 (13%) |
| **Neutrophils, median (IQR)** | 6.2 (4.3, 8.8) | 6.9 (4.3, 10.2) | 5.2 (3.4, 8.0) | 8.1 (5.8, 10.9) | 7.8 (5.2, 11.7) |
| **Not initially measured** | 54 (17%) | 93 (21%) | 72 (18%) | 7 (6%) | 9 (12%) |
| **Lymphocytes, median (IQR)** | 0.9 (0.7, 1.3) | 0.8 (0.6, 1.2) | 1.1 (0.8, 1.6) | 1.0 (0.7, 1.3) | 0.8 (0.6, 1.2) |
| **Not initially measured** | 54 (17%) | 93 (21%) | 72 (18%) | 8 (7%) | 10 (13%) |
| **Sodium , median (IQR)** | 137 (134, 139) | 137 (134, 141) | 136 (134, 138) | 136 (133, 138) | 137 (134, 139) |
| **Not initially measured** | 49 (15%) | 84 (19%) | 67 (17%) | 5 (4%) | 9 (12%) |
| **Potassium , median (IQR)** | 4 (4, 4) | 4 (4, 4) | 4 (4, 4) | 4 (4, 4) | 4 (4, 4) |
| **Not initially measured** | 49 (15%) | 84 (19%) | 67 (17%) | 5 (4%) | 9 (12%) |
| **Urea , median (IQR)** | 8 (6, 11) | 9 (7, 14) | 5 (4, 8) | 6 (5, 9) | 9 (6, 12) |
| **Not initially measured** | 49 (15%) | 84 (19%) | 67 (17%) | 5 (4%) | 9 (12%) |
| **Creatinine, median (IQR)** | 88 (66, 120) | 96 (72, 151) | 74 (60, 92) | 79 (60, 106) | 99 (74, 162) |
| **Not initially measured** | 49 (15%) | 84 (19%) | 67 (17%) | 5 (4%) | 9 (12%) |
| **O2 saturations, median (IQR)** | 96 (95, 97) | 96 (94, 97) | 96 (95, 97) | 95 (93, 96) | 95 (92, 97) |
| **Not initially measured** | 3 (1%) | 8 (2%) | 23 (6%) | 0 (0%) | 3 (4%) |
| **FiO2 , median (IQR)** | 28.0 (24.4, 33.7) | 29.5 (25.6, 43.2) | 26.9 (21.0, 30.4) | 39.0 (30.2, 52.7) | 41.4 (29.7, 56.3) |
| **Not initially measured** | 92 (28%) | 130 (29%) | 130 (33%) | 7 (6%) | 16 (21%) |
| **Respiratory Rate , median (IQR)** | 19.1 (18.1, 20.8) | 19.7 (18.1, 22.5) | 19.1 (17.9, 20.8) | 23.5 (19.3, 27.9) | 20.3 (18.2, 25.6) |
| **Not initially measured** | 3 (1%) | 8 (2%) | 22 (6%) | 9 (8%) | 4 (5%) |
| **Heart rate , median (IQR)** | 83.0 (74.2, 91.0) | 86.2 (75.9, 95.6) | 85.7 (76.1, 93.6) | 91.0 (81.4, 99.6) | 89.4 (78.1, 98.2) |
| **Not initially measured** | 3 (1%) | 8 (2%) | 23 (6%) | 9 (8%) | 4 (5%) |
| **Systolic Blood Pressure, median (IQR)** | 131 (118, 146) | 128 (117, 140) | 127 (118, 137) | 126 (117, 142) | 127 (115, 141) |
| **Not initially measured** | 3 (1%) | 8 (2%) | 29 (7%) | 9 (8%) | 4 (5%) |
| **Diastolic Blood Pressure, median (IQR)** | 70 (64, 76) | 68 (63, 74) | 71 (66, 76) | 70 (66, 78) | 70 (62, 76) |
| **Not initially measured** | 3 (1%) | 8 (2%) | 29 (7%) | 9 (8%) | 4 (5%) |
| **Temperature, median (IQR)** | 37 (36, 37) | 37 (36, 37) | 37 (37, 37) | 37 (37, 38) | 37 (36, 37) |
| **Not initially measured** | 3 (1%) | 9 (2%) | 22 (6%) | 9 (8%) | 4 (5%) |

**Table 3.** Risk prediction models for next day escalation or death amongst patients eligible for escalation, and for next day mortality amongst patients not eligible for escalation 21 February 2020 until 30 June 2020 (derivation cohort)

| Hazard ratios<br>(95% Confidence<br>intervals) | Patients not for<br>escalation: Next day<br>death | Patients for<br>escalation:<br>Next day ICU<br>admission or death | Bootstrapped samples<br>in which covariate<br>selected by AIC for next<br>day escalation or death |
| --- | --- | --- | --- |
| Daily mean<br>Haemoglobin | 0.99 (0.99-1.00) |  |  |
| Within day change in<br>Haemoglobin |  | 0.95 (0.92-0.98) | 84% |
| Lagged change in daily<br>mean Haemoglobin |  | 0.98 (0.97-0.99) | 45% |
| Log(mean daily<br>Neutrophils) | 1.45 (1.22-1.67) | 1.88 (1.47-2.28) | 83% |
| log(daily mean<br>Lymphocyte count) | 0.80 (0.62-0.98) |  |  |
| lagged daily change in<br>mean Lymphocyte<br>count | 0.67 (0.28-1.06) |  |  |
| log(Platelet count) | 0.69 (0.46-0.91) | 0.42 (0 - 0.95) | 79% |
| Daily mean Sodium | 1.03 (1.02-1.05) | 0.95 (0.89-1.00) |  |
| Within day change in<br>Sodium |  | 1.13 (1.01-1.24) | 50% |
| Lagged change in daily<br>mean Potassium | 1.61 (1.40-1.82) | 2.09 (1.71-2.48) | 65% |
| log(daily mean Urea) | 2.16 (1.81-2.50) |  |  |
| Within day change in<br>Urea | 1.07 (1.03-1.12) |  |  |
| Lagged change in daily<br>mean Urea | 0.93 (0.88-0.97) | 0.94 (0.88-1.01) | 23% |
| log(mean daily<br>Creatinine) | 0.65 (0.31-0.98) |  |  |
| Within day change in<br>Creatinine |  | 1.01 (1.00-1.02) | 35% |
| Highest daily<br>Temperature | 0.72 (0.54-0.89) | 1.60 (1.33-1.87) | 99% |
| Highest respiratory rate | 1.09 (1.07-1.11) | 1.10 (1.07-1.13) | 88% |
| Within day change in<br>respiratory rate | 1.04 (1.01-1.06) |  |  |
| Lagged daily change in<br>highest respiratory rate | 0.97 (0.95-0.98) | 0.96 (0.93-0.99) | 60% |
| Highest daily heart rate | 1.01 (1.00-1.02) |  |  |
| Within day change in<br>heart rate | 1.01 (1.00-1.02) | 1.03 (1.01-1.05) | 83% |
| Lagged change in<br>highest daily heart rate |  | 1.02 (1.01-1.03) | 48% |
| Lowest daily diastolic<br>blood pressure | 1.01 (1.00-1.02) |  |  |
| Highest daily FiO2 (%) | 1.03 (1.02-1.03) | 1.04 (1.03-1.05) | 100% |
| Daily lowest oxygen saturation (%) | 0.98 (0.97-0.99) |  |  |
| Age on admission (years) |  | 1.21 (1.11-1.30) | 61% |
| Age<sup>2</sup> on admission (years) |  | 1.00 (1.00-1.00) | 60% |
| BMI < 20 | 1.81 (1.51-2.11) |  |  |
| Current Smoker | 0.44 (0-0.88) |  |  |
| South Asian ethnic group | 3.33 (2.49-4.17) |  |  |
| Total observed patient days | 9,395 | 6,066 |  |
| Log Likelihood | -1,280.65 | -344.59 |  |
| LR Test | 619.57 (df = 23) | 238.90 (df = 17) |  |

On an individual level the performance of this model for predicting next day escalation or death outcome improved from the admission only model with an overall concordance of 0.90 (95% CI 0.86-0.93) in the whole dataset, that only reduced to a median 0.87 (IQR 0.85-0.90) with cross validation using bootstrapped samples, and had high daily discrimination when assessed across the follow up time as assessed with leave one out cross validation (supplementary figure S4). Restricting the population to just those that had SARS-CoV-2 PCR positive test did not alter the bootstrapped discrimination (0.87 (IQR 0.84-0.89)).

Falling haemoglobin, rising neutrophil count, low platelets, worse renal function, low sodium, rising electrolytes/creatinine, higher temperature, high but falling respiratory rate, rising heart rate, and higher oxygen requirement, and age were associated with next day ICU admission or death as might be expected. The addition of further demographics (e.g. ethnicity, and gender), co-morbidity and life style factors did not predict next day deterioration independently of these included covariates. Consequently, although Black and Asian ethnic groups were initially protective within this cohort after adjusting for the other covariates, but with small numbers this result was not robust and was not selected in the final model.

### Patients ineligible for escalation in 1^st^ wave: Predicting next day mortality

As a comparison to the previous analysis, a model was built predicting next day mortality in those patients not eligible for escalation to ICU. The model is shown in the first column in table 3, and, it shows a similar pattern in hazard ratios for most of the shared predictors. The model’s leave one out cross validation discrimination was 0.86 (0.85 - 0.87), and remained high throughout the follow up in leave one out cross validation (figure S5). Baseline survival plots are shown in supplementary figure S6.

### Calibration and Comparison with existing scores in first wave

To assess calibration, the proportion of patients with next day escalation was calculated for each quintile of magnitude of the linear predictor in those with outcomes. This was calculated using the centred means of the covariates as described in the equations in table S2, and(boundaries for the linear predictor set at values of −6.5, 2.1, 3.4, 4.4, 5.3, and 7.5 using the quintiles of the score in patients with events in the derivation cohort.. This was compared to the proportion in quintiles calculated from NEWS2 (boundaries at values of 0, 1, 2, 3, 5, and 14) and the ISARIC 4C mortality score (boundaries at values of 0, 6.0, 8.0, 10.2, 13.0, and 20.0). Figure 2a shows that our derived score had better calibration for next day escalation among those patients who were eligible. The corresponding discrimination for the ISARIC 4C mortality score was C statistic = 0.58 and for the NEWS2 score C statistic = 0.80 in the derivation cohort.

**Figure 2.**
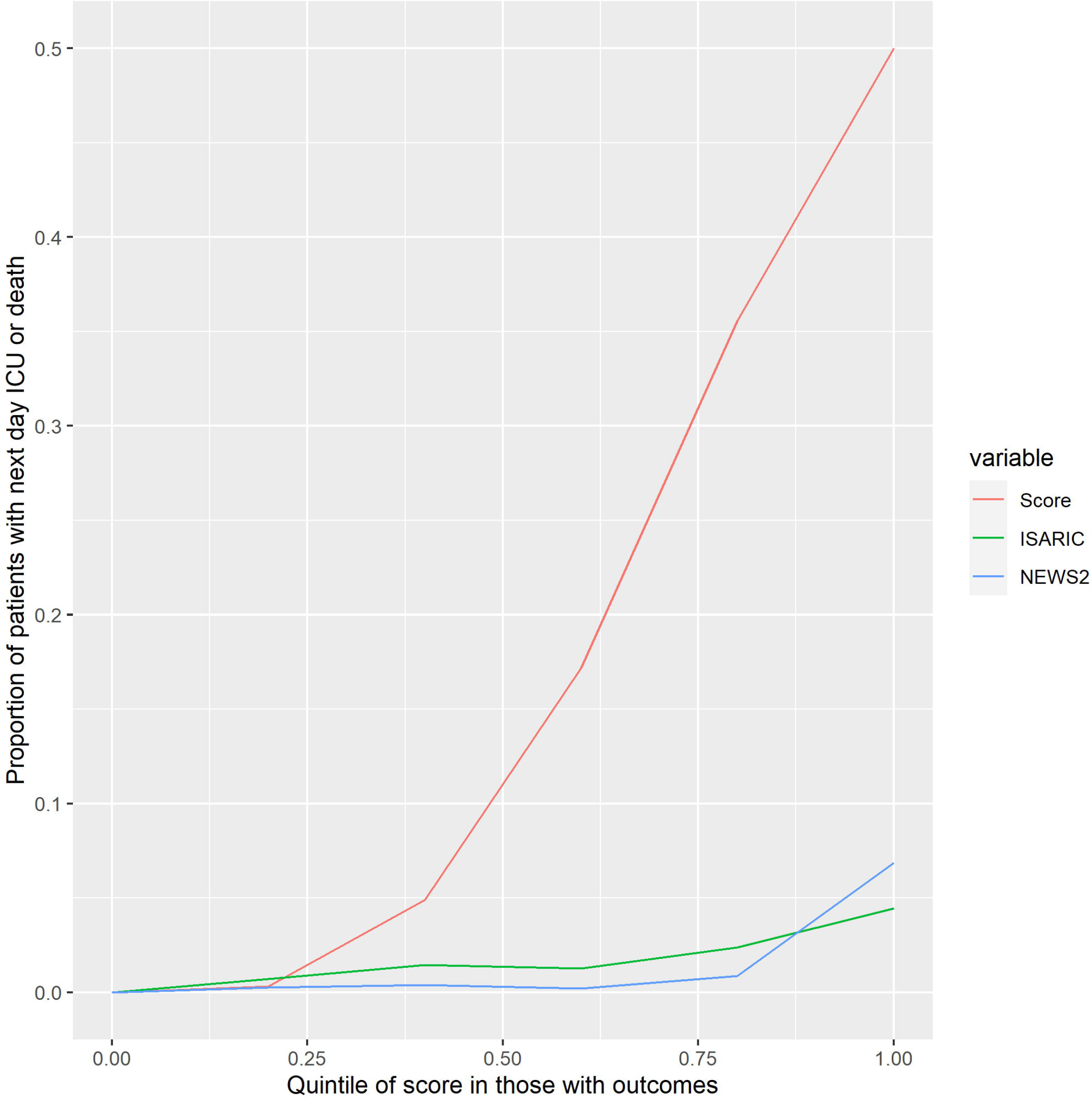
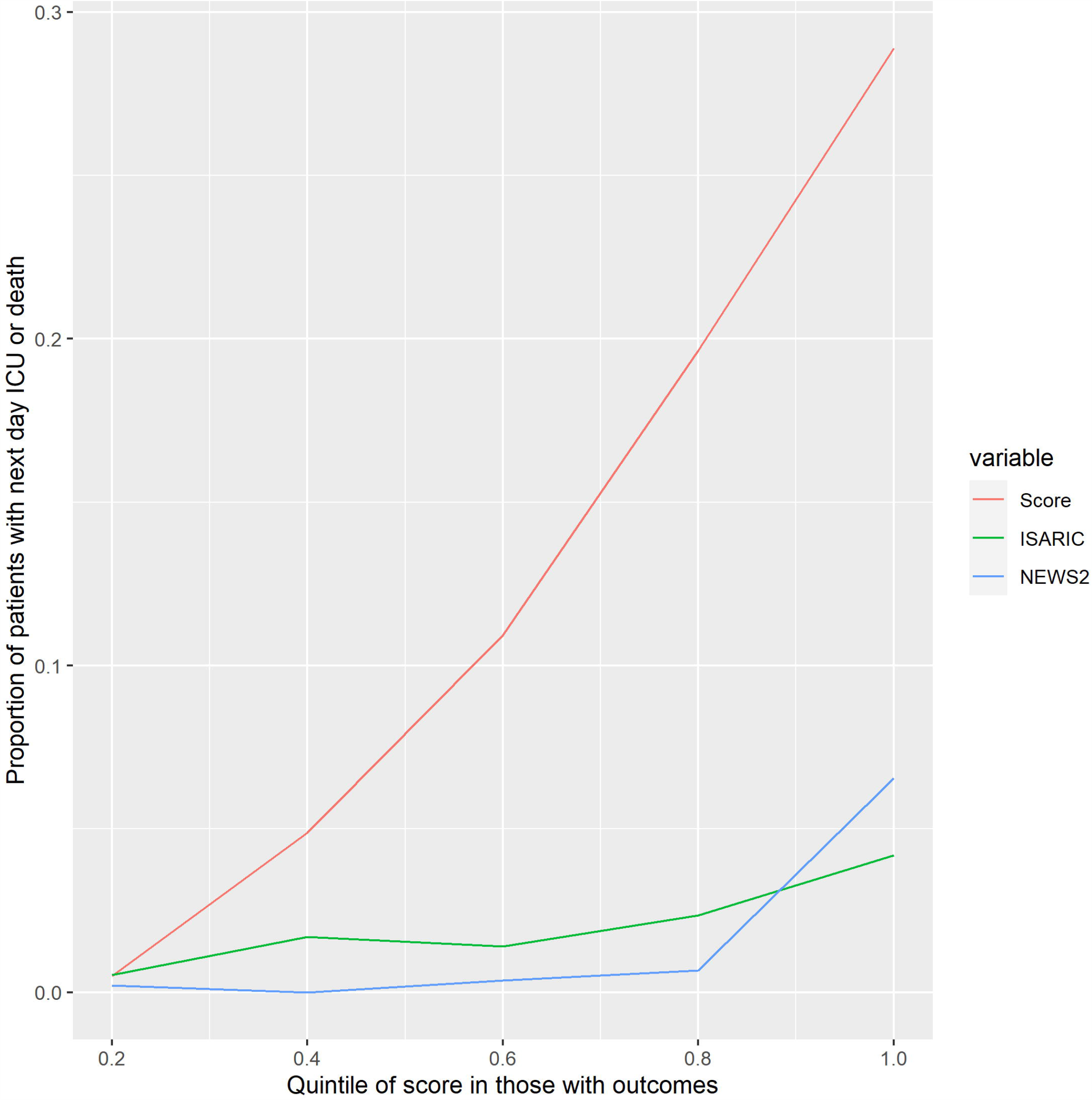
Plot of the proportion of patients escalated the following day who were eligible within each quintile of; calculated score (derived within this study), highest daily NEWS2, and ISARIC 4C mortality score. 2 (a) Calculated with leave one out cross validation in derivation cohort 2 (b) Calculated within the validation cohort

In addition, when compared over time, the magnitude of the score for eligible inpatients tracked the observed number of escalations (figure S8a), and the score predicting next day mortality for ineligible patients tracked the observed mortality (figure S9a).

Taking some of the key predictors derived in this study, only 0.02% of eligible patient days with an FiO2 <30%, Temperature <38°C, Neutrophil count under 10 x 10^9^/l, and normal creatinine, had a subsequent day escalation. In contrast, 44% of patients with an FiO2 >30%, Temperature >=38°C, Neutrophil count >=10 x 10^9^/l, and a raised creatinine had a next day escalation.

### Second Wave Validation

The score derived in the first wave was applied to all patients admitted to NUH with SARS-CoV-2 who were confirmed clinically or by PCR from July to end November 2020. The discrimination of the score for patients eligible for escalation remained similar to the derivation cohort with a concordance of 0.88 (95% CI 0.81 −0.95). The corresponding discriminations were also improved in the validation cohort compared to the derivation cohort for the ISARIC-4C mortality score (C statistic = 0.70 (95% CI 0.63-0.77)) and for the NEWS2 score (C statistic = 0.89 (95% CI 0.82 – 0.96).

To assess calibration, the proportion of patients with next day escalation was calculated for each quintile of magnitude of the linear predictor in those with outcomes using the boundaries from the derivation cohort (Figure 2b). Whilst maintaining its discrimination this shows the score overestimated next day escalation in the validation cohort compared to the derivation cohort with a calibration slope of 0.55 (figure S7).

For patients ineligible for escalation the model predicting next day mortality had a discrimination of 0.86 (95% CI 0.82-0.89) and a calibration slope of 0.78. In comparison the discrimination of the daily NEWS score for next day mortality was 0.82 (95% CI 0.78-0.85), and the ISARIC-4C score 0.76 (95% CI 0.72-0.79).

When compared over time, the magnitude of the score for eligible inpatients tracked the observed number of escalations (figure S8b), and the score predicting next day mortality for ineligible patients tracked the observed mortality (figure S9b) at a higher score value than in the derivation cohort.

## Discussion

### Main findings

This study describes the characteristics and clinical outcomes of a complete cohort of patients admitted to a single hospital in England throughout the SARS-CoV-2 pandemic from February to November 2020. In patients eligible for escalation of care we found that several factors were associated with prognosis both at baseline and throughout the course of their illness. Our model incorporating daily clinical and laboratory measurements in addition to baseline characteristics was able to predict the need for escalation of care, death or survival without escalation of care with good precision throughout the admission. The validation showed excellent discrimination, but that the score needed to be recalibrated as at thresholds from the derivation cohort it over predicted death and escalation. This is likely to reflect the change in demographics and clinical practice between the first and second UK wave, given changes in escalation practice[11,12] and introduction of the use of steroids in patient treatment[13,14]. Our results if proven valid in other populations suggest that a score such as ours could provide a warning of the need for escalation of care in SARS-CoV-2 patients. In addition by integration of scoring across an admitted patient cohort in a hospital we can potentially predict demand for ICU resources.

### Strengths and weaknesses

Our study included all patients who were admitted to a single hospital serving the population of a city throughout an eight month period of 2020 and so should be widely generalisable to similar populations elsewhere. Unlike many other reports we have included all patients both clinically diagnosed and PCR confirmed meaning that our findings should be generalisable where such clinical decisions have been taken. Our sensitivity analysis shows that our model performed better when restricted to those PCR confirmed, and table S1 suggests those patients with more severe disease were more likely to have a positive test. This is likely to reflect the longer period of time in hospital these patients had during which they would be retested and will be less of an issue in more recent data with more rapid test processing. Through our use of electronic patient record systems, we had access to comprehensive sociodemographic, clinical and laboratory variables including all measurements recorded electronically through the patient’s admission. We also had available complete follow up for escalation of care, death (including out of hospital death) or discharge from hospital for 30 days from admission and importantly, therefore, have little bias due to missing data, loss to follow up or other common biases of observational cohorts. However it must be recognised that though this richness and uniformity of data is a strength, it is gained at the cost of limiting our analyses to one city and the decisions of one cohort of clinicians. This leads to questions regarding generalisability which can only be answered by external validation. However the diverse population of Nottingham and the standardisation of care across the NHS suggest we believe that our findings will be replicable. To increase the validity of our findings our modelling of the risk accounted for eligibility for escalation of care in defining the relevant outcomes.

### Interpretation

Our report is best compared to those other large population-based studies from single cities or regions around the world that have reported their experience through the SARS-CoV-2 pandemic[15–22] and the relevant UK studies[23–25]. The distribution of sociodemographic risk factors and their association with poor prognosis with respect to age and sex are similar to these studies. Differences are apparent mainly in relation to ethnicity which is unsurprising given the different populations included elsewhere in the world[15–19]. Our risk prediction model is unique in that it has utilised longitudinal daily clinical and laboratory measures, alongside baseline characteristics, to estimate the daily need for escalation of care or death. In that respect, we cannot compare it directly to other published risk models but in relation to those derived within UK populations, it performs better[5,8,25] and for reasons stated above is at low risk of bias. In particular, compared to the robustly developed ISCARIC 4C mortality prediction score[5] our model performs better on a daily basis – showing the value of incorporating repeated measurements of clinical observations and blood results into the prediction of prognosis for patients admitted to hospital with SARS-CoV-2.

## Conclusions

We have shown that incorporating daily measurements of clinical observations and blood tests improves the accuracy of both the prediction of prognosis and of resource demand in secondary care patients with SARS-CoV-2. Clinical application of such a dynamic score could be used to prompt clinical review to ensure timely escalation of care, and to predict the need to increase or repurpose critical care capacity at an operational level in hospitals.

## Supporting information

Tripod checklist

