## Supplementary tables and figures for "Predicting the need for escalation of care or death from repeated daily clinical observations and laboratory results in patients with SARS-CoV-2 during 2020: a retrospective population-based cohort study from the United Kingdom"

Table S1. Sociodemographic and other characteristics on admission of the cohort who were admitted to hospital with confirmed SARS-COV-2 diagnosis 21 February 2020 until 30 June 2020

|  | Admission: Clinical diagnosis only | Admission: PCR positive |
| --- | --- | --- |
| N | 388 | 986 |
| Age (years) median (IQR) | 76 (61, 85) | 76 (60, 84) |
| Male | 191 (49%) | 537 (54%) |
| Other or not stated ethnic group | 59 (15%) | 188 (19%) |
| Black/Mixed ethnic group | 9 (2%) | 43 (4%) |
| Indian/Pakistani ethnic group | 10 (3%) | 39 (4%) |
| White ethnic group | 310 (80%) | 716 (73%) |
| 30 day re-admission | 141 (36%) | 323 (33%) |
| 30 day mortality | 85 (22%) | 283 (29%) |
| Died out of hospital | 13 (3%) | 26 (3%) |
| 30 day ICU admission | 22 (6%) | 129 (13%) |
| Length of stay, days (IQR) | 8 (4, 13) | 8 (3, 17) |
| For escalation/CPR | 156 (40%) | 437 (44%) |
| NEWS2, median (IQR) | 4 (2, 6) | 3 (2, 6) |
| ISARIC-4C, median (IQR) | 10 (7, 12) | 10 (6, 12) |
| BMI <20, n, % | 70 (18%) | 149 (15%) |
| BMI >30,n, % | 92 (24%) | 291 (30%) |
| Smoking, n, % | 70 (18%) | 81 (8%) |
| Vaping, n, % | 23 (6%) | 36 (4%) |
| Alcohol risk, n, % | 54 (14%) | 145 (15%) |
| Charlson Index, median (IQR) | 2 (1, 4) | 1 (0, 3) |

Table S2. Calculation of linear predictors for models used for validation in second wave

Linear predictor for next day escalation/death model for patients eligible for escalation =

(-0.044579534 * (Within day change in_Haemoglobin +0.2869029)) +

(-0.021847427 * (Lagged change in daily_mean Haemoglobin - 6.916713)) +

(0.645442813 * (log daily_mean Neutrophils - 1.575847 )) +

(-0.875495011 * (log daily_mean Platelet Count - 5.549459)) +

(-0.058933854 * (Daily_mean Sodium - 135.7257 )) +

(0.117616898 * ( Within day change in mean Sodium - 0.01032357 )) +

(0.760791874 * (Lagged change in daily mean Potassium - 0.2330512)) +

(0.066315384 * (Lagged change in daily mean Urea -0.3525179)) +

(0.009527933 * (Within day change in mean Creatinine +0.527812)) +

(0.491759315 * (Highest daily Temperature - 36.81334)) +

(0.092938489	* (Highest daily respiratory rate - 19.66502 )) +

(-0.039391416	* (Lagged change in highest daily respiratory rate - 1.231279 )) +

0.027517011 * (Within day change in highest heart rate - 0.09753467 )) +

(0.023452761 * (Lagged daily change in highest heart rate - 5.185362 )) +

(0.037472868 * (Highest daily FiO2 - 26.12712)) +

(0.189021008 * (Age on admission - 62.76595)) +

(-0.001531570 * (Age on admission^2 - 4294.754))

Linear predictor for next day death model for patients ineligible for escalation =

(-0.008398623	* (Daily mean Haemoglobin - 111.9086)) +

(0.349750124	* (log daily_mean Neutrophils - 1.690934)) +

( -0.214921615	* (log daily_mean Lymphocytes - -0.01981265)) +

( -0.408232993	* (Lagged change in daily_mean Lymphocytes - 0.07047588 )) +

(-0.380651164 * (Log daily_mean Platelet Count - 5.414716)) +

(0.029376232 * (Daily_mean Sodium - 137.5975 )) +

(0.476896137 * (Lagged change in daily_mean Potassium - 0.206123)) +

(0.794877867 * (Log daily_mean Urea - 2.034817)) +

( 0.078782897 * (Within day change in_Urea -	-0.07661035)) +

(-0.080712669	* (Daily mean Urea -	0.4939551)) +

(-0.408693227 * (Log daily_mean Creatinine -	4.492894)) +

(-0.339623523 * (Highest daily temperature - 36.96255)) +

(0.084128451 * (Highest daily respiratory rate - 20.77068 )) +

(0.036964020 * (Within day change in respiratory rate - 0.03683924)) +

(-0.034325467 * (Lagged daily change in highest respiratory - 1.129918)) +

(0.011032458 * (Highest daily heart rate - 90.03662)) +

(0.010107580 * (Within day change in heart rate +0.001961853))	+

(0.014596208 * (Lowest daily diastolic blood pressure - 60.66376)) +

(0.025323205 * (Highest daily FiO2 - 30.36593)) +

(-0.018695667 * (Lowest daily oxygen saturation - 93.50075)) +

(0.647398803 * ((BMI < 20) - 0.1431063)) +

(-0.857695694	* ((Is a Current Smoker) - 0.1297003)) +

(1.255664474 * (Ethnicity South Asian - 0.01177112))

Figure S1. Cohort from first suspected date between 21 February 2020 until 30 June 2020

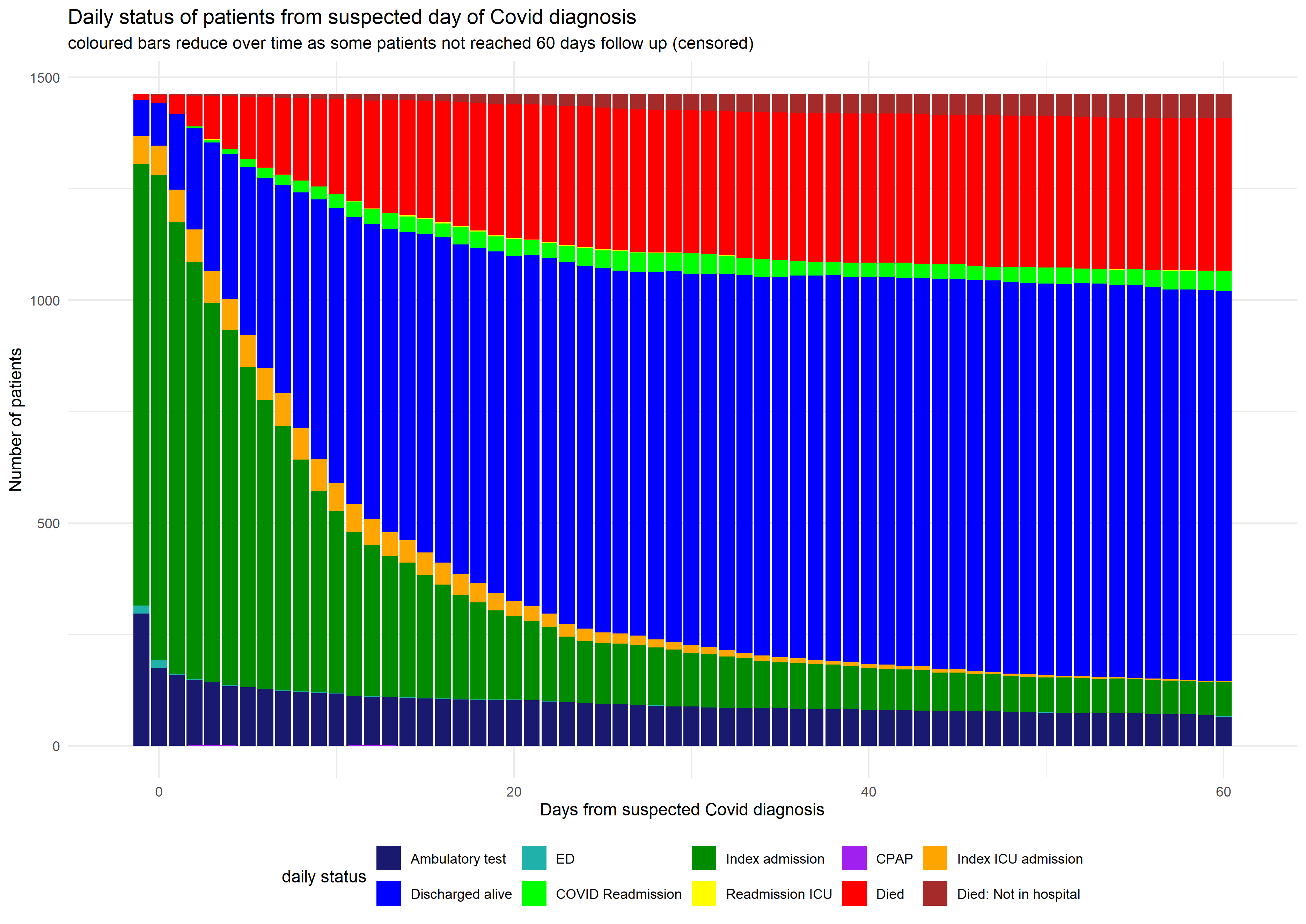

Figure S2a. Flow chart of all suspected SARS-COV-2 patients in the derivation cohort until 30th June 2020

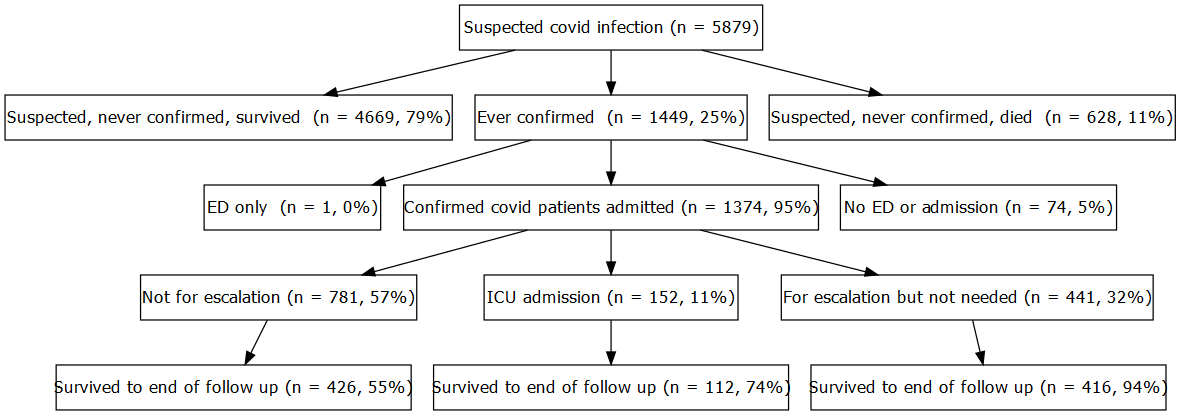

Figure S2b. Flow chart of all suspected SARS-COV-2 patients in the validation cohort from 1st July 2020

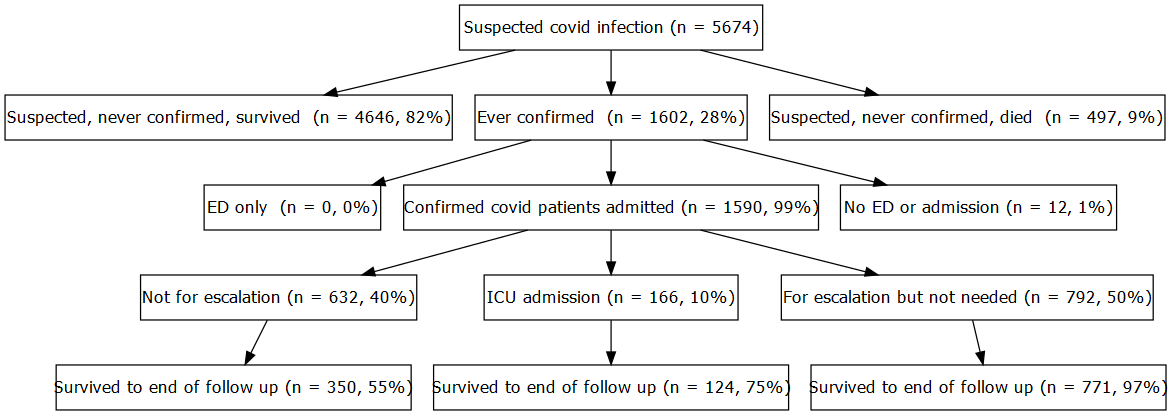

Figure S2c. Flow chart of patients with both blood and observation measurements before outcome observed in derivation cohort

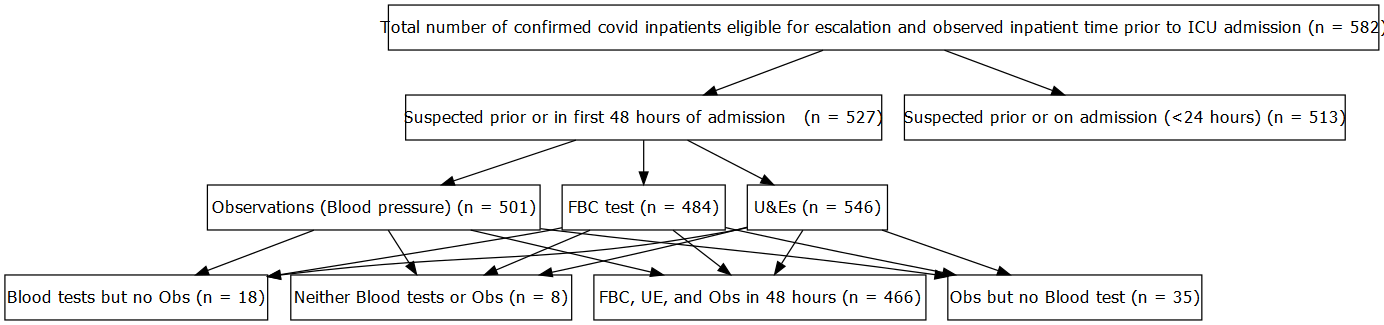

Fig S3 Longitudinal smoothed means of selected observations and blood tests by observed worst outcome

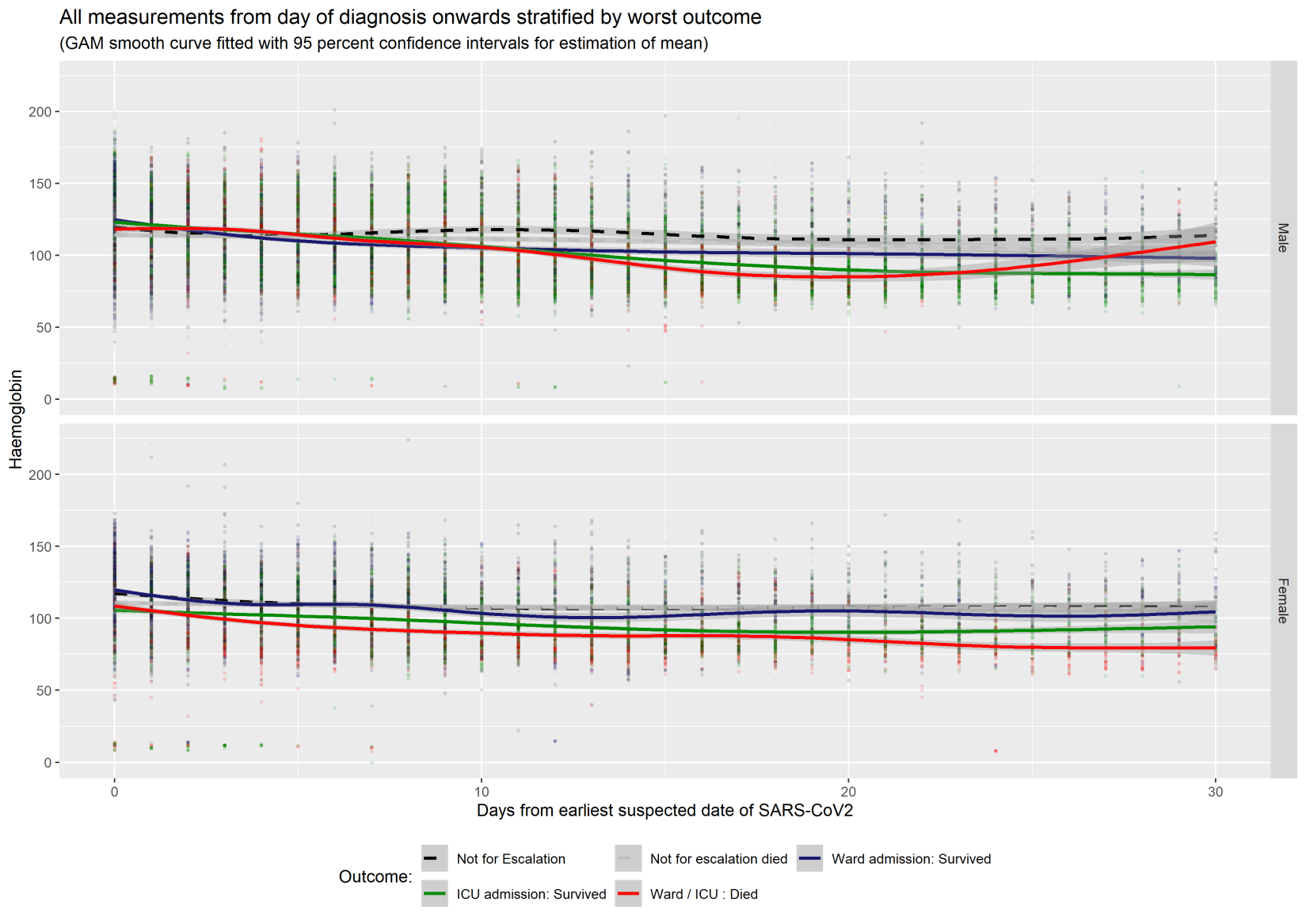

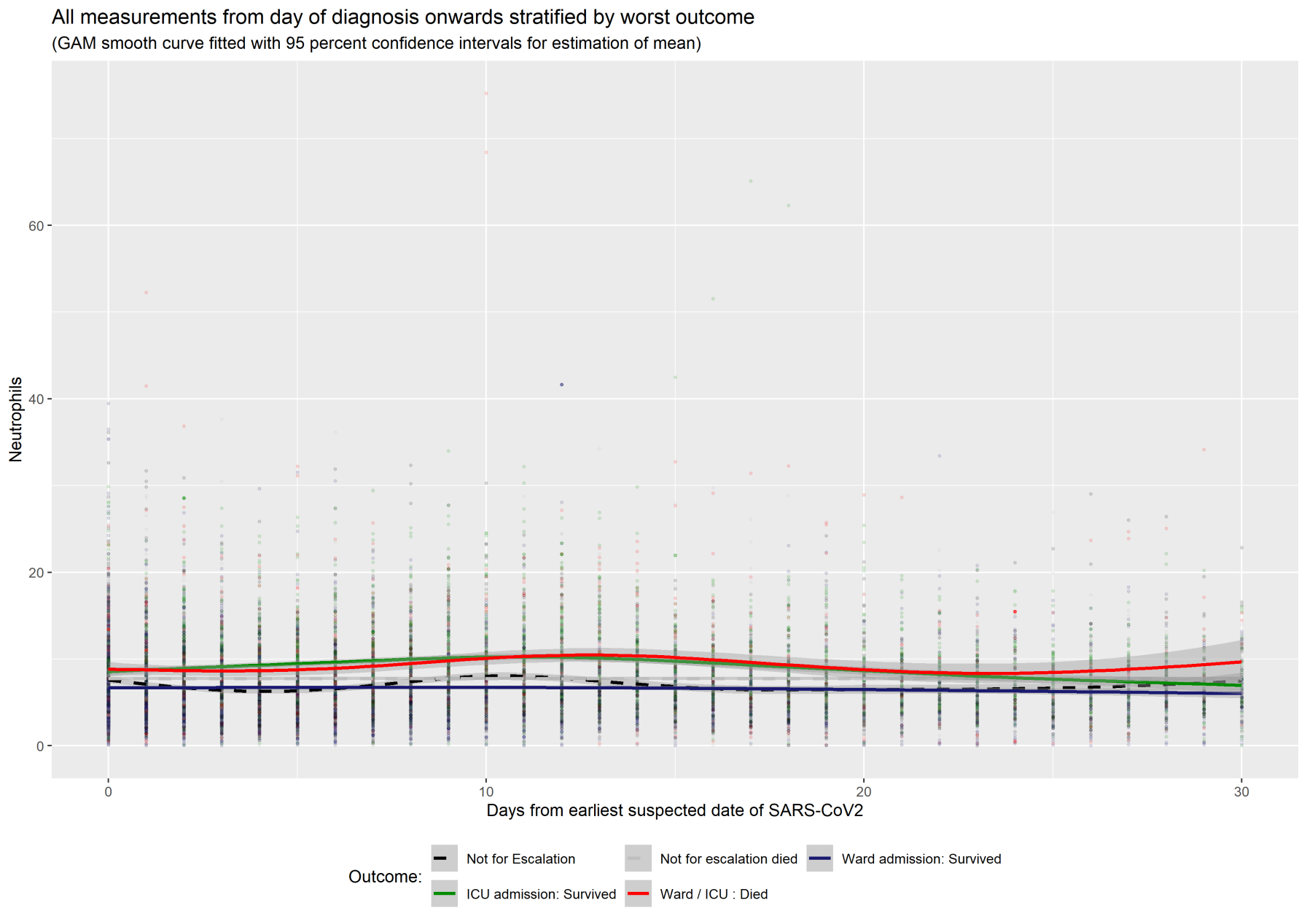

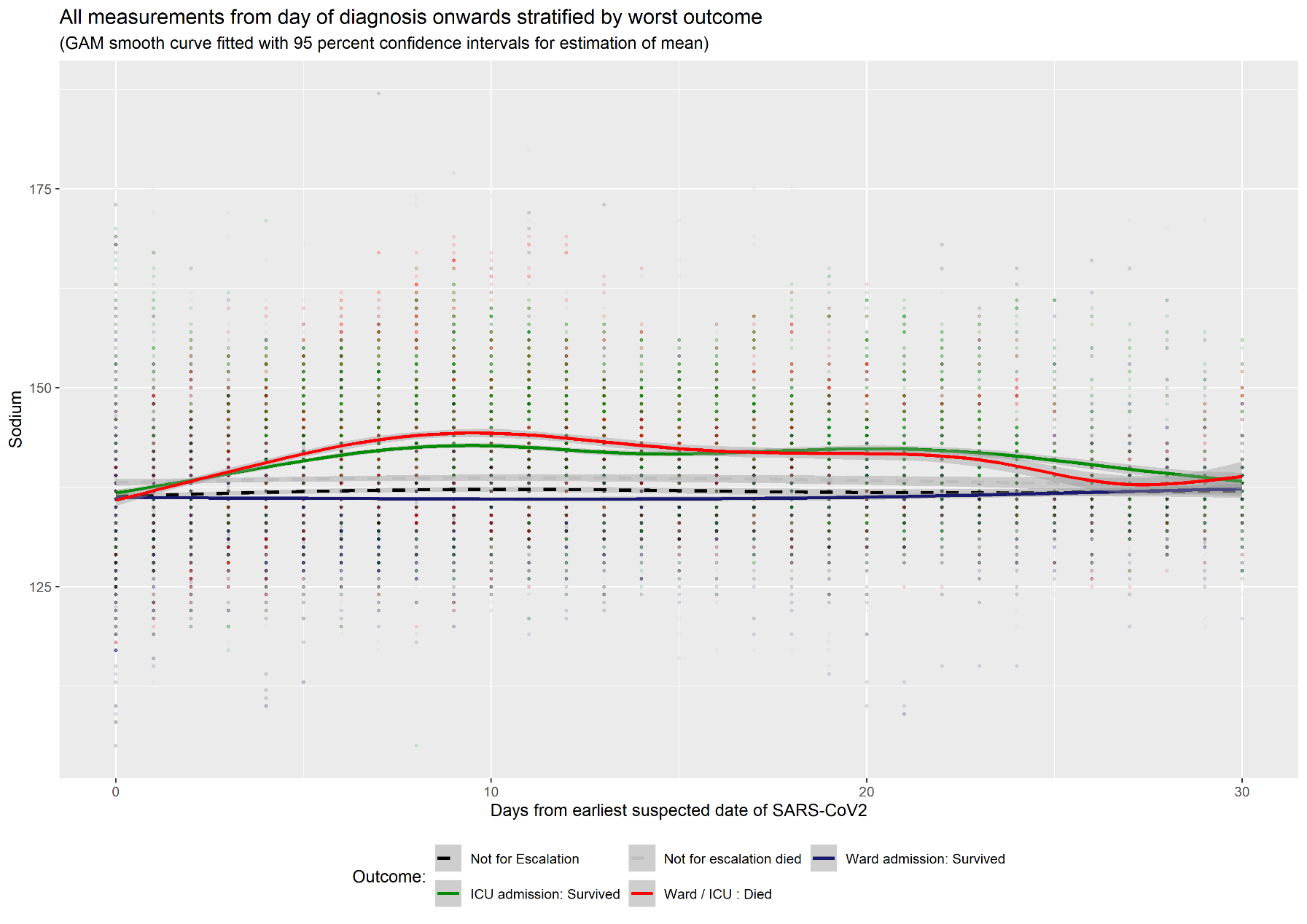

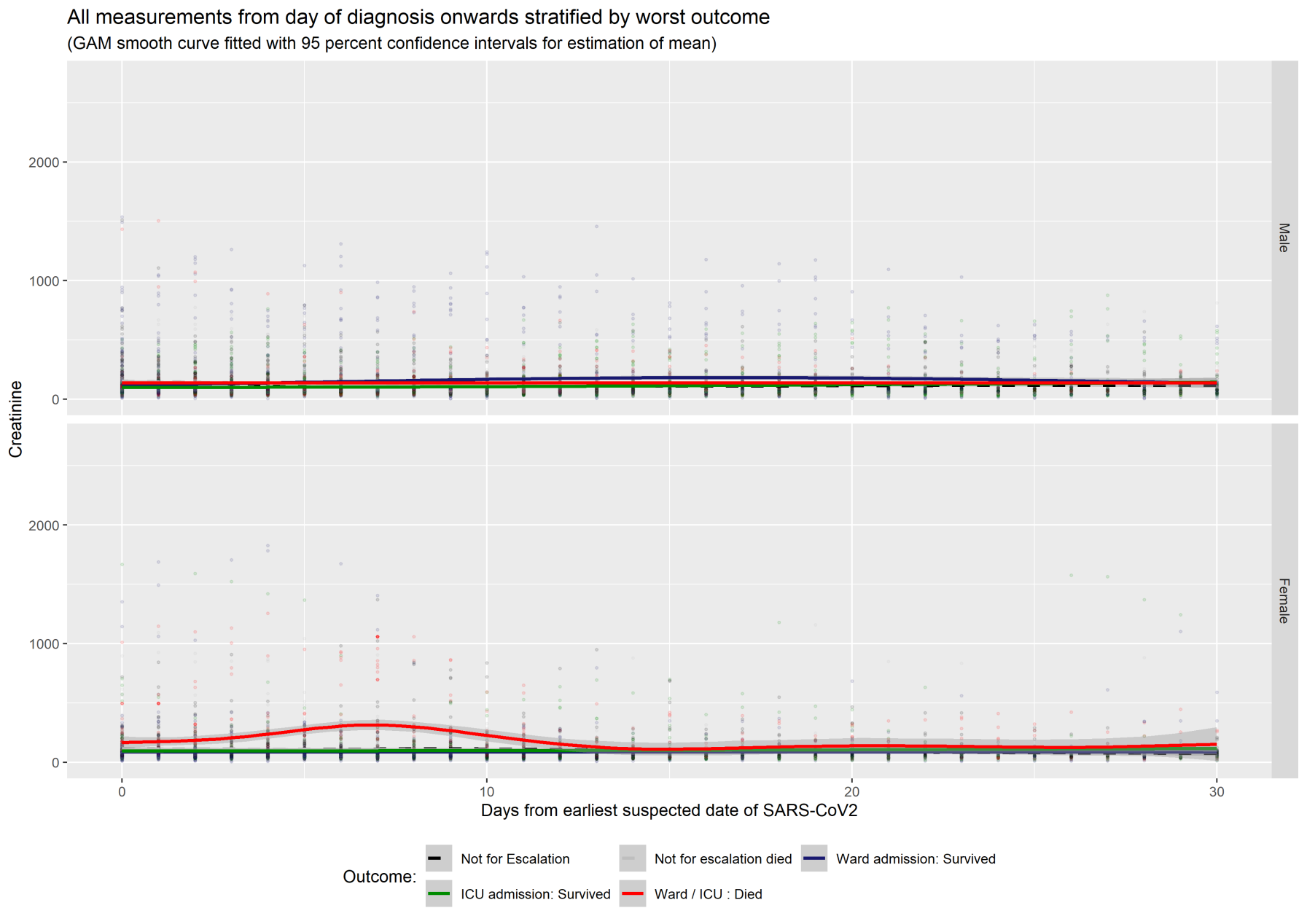

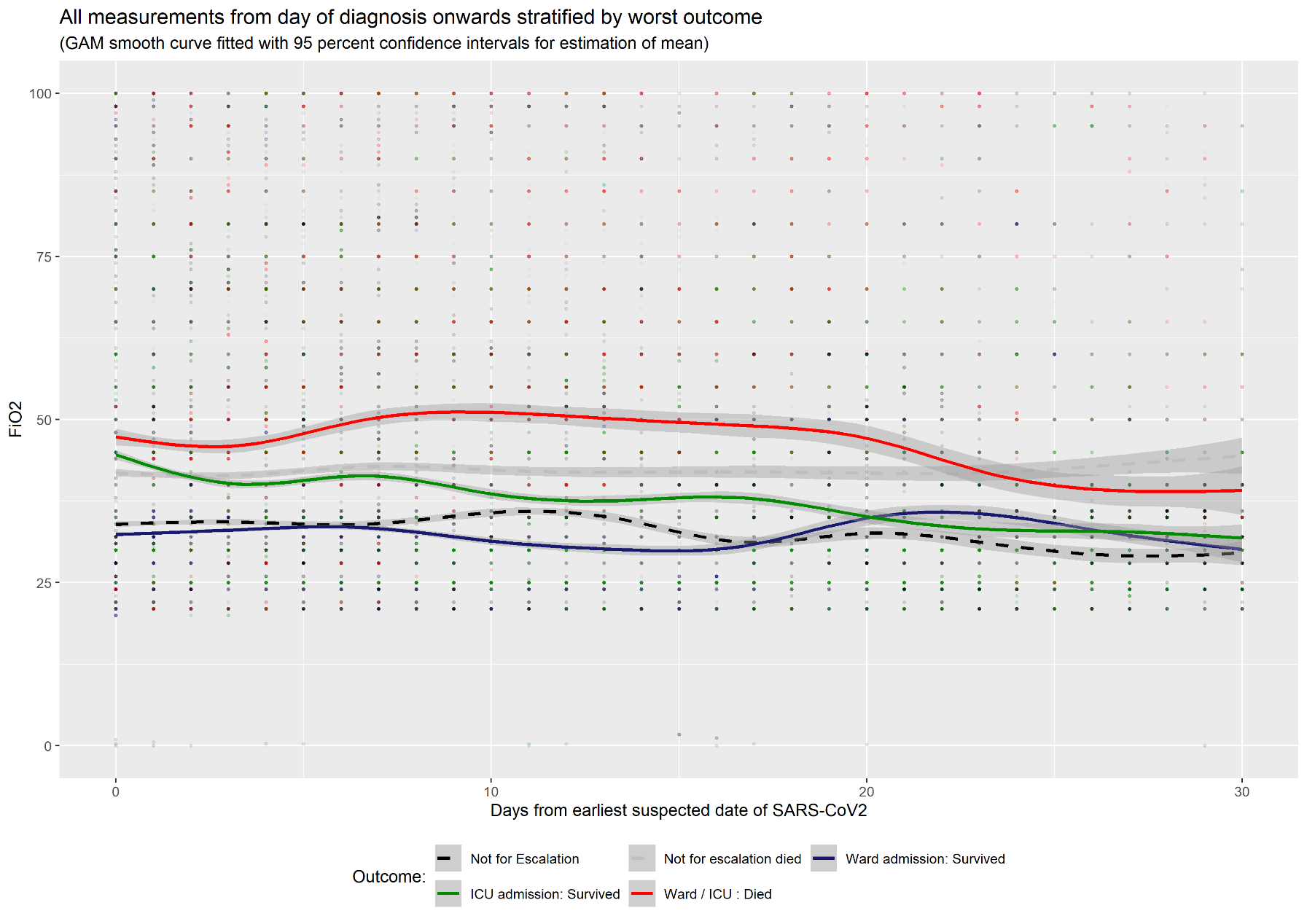

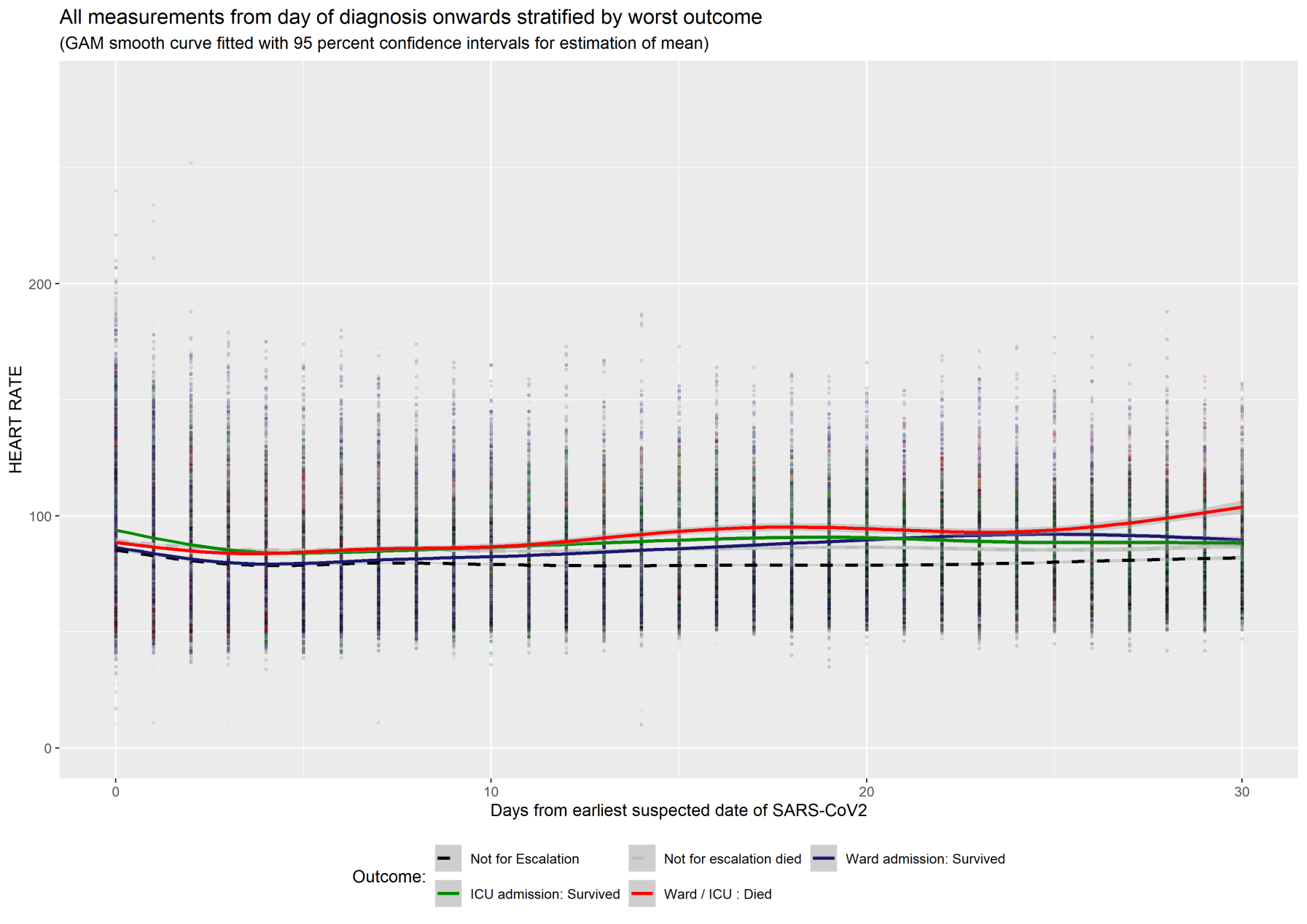

Figure S4. ROC curves and AUC for daily prediction of next day escalation from time varying model by day of disease course within patients eligible for escalation 21 February 2020 until 30 June 2020 with leave one out cross validation

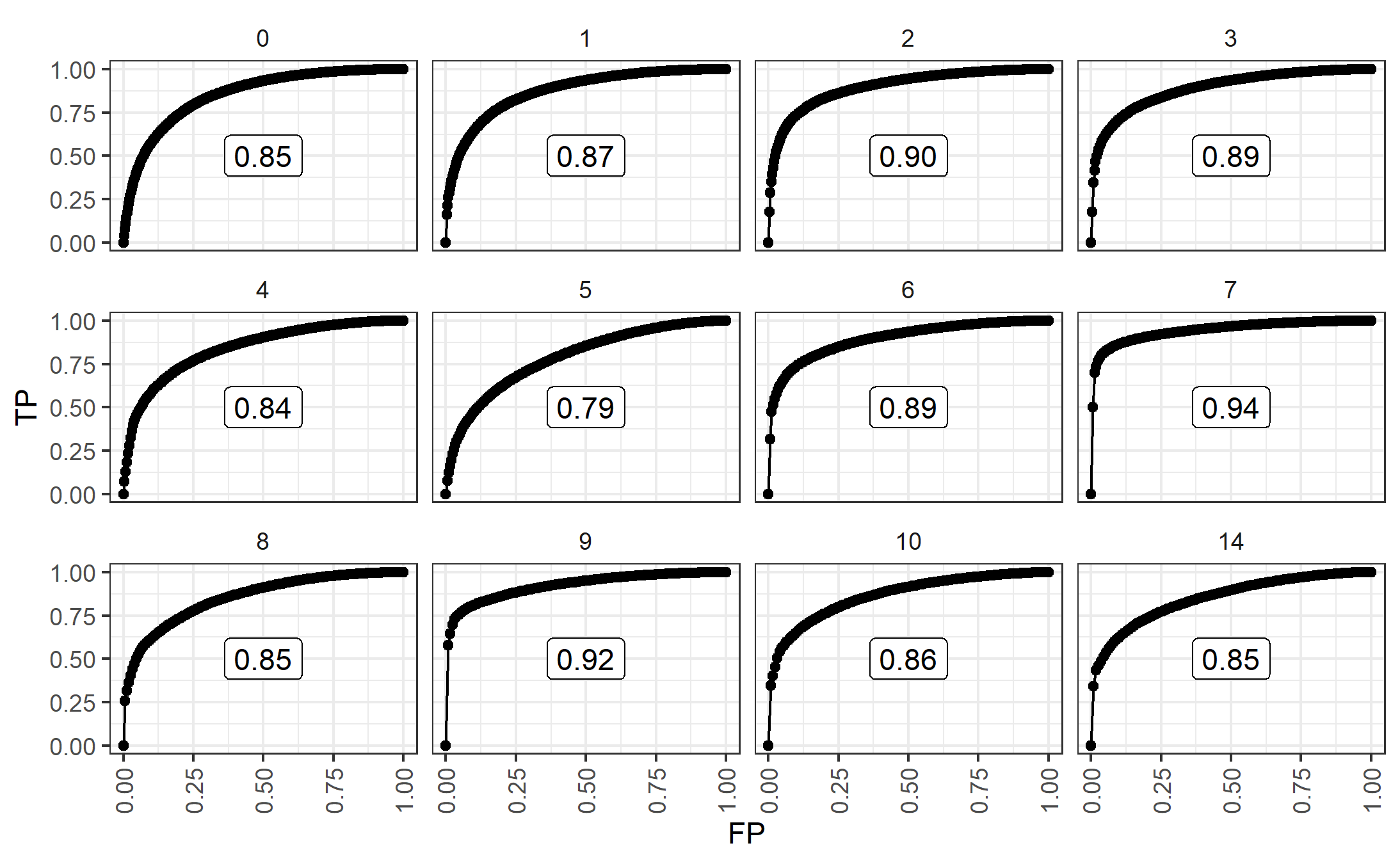

**Figure S5:**

ROC curves and AUC for daily prediction of next day mortality by day of disease course for patients ineligible for escalation for patients 21 February 2020 until 30 June 2020 with leave one out cross validation

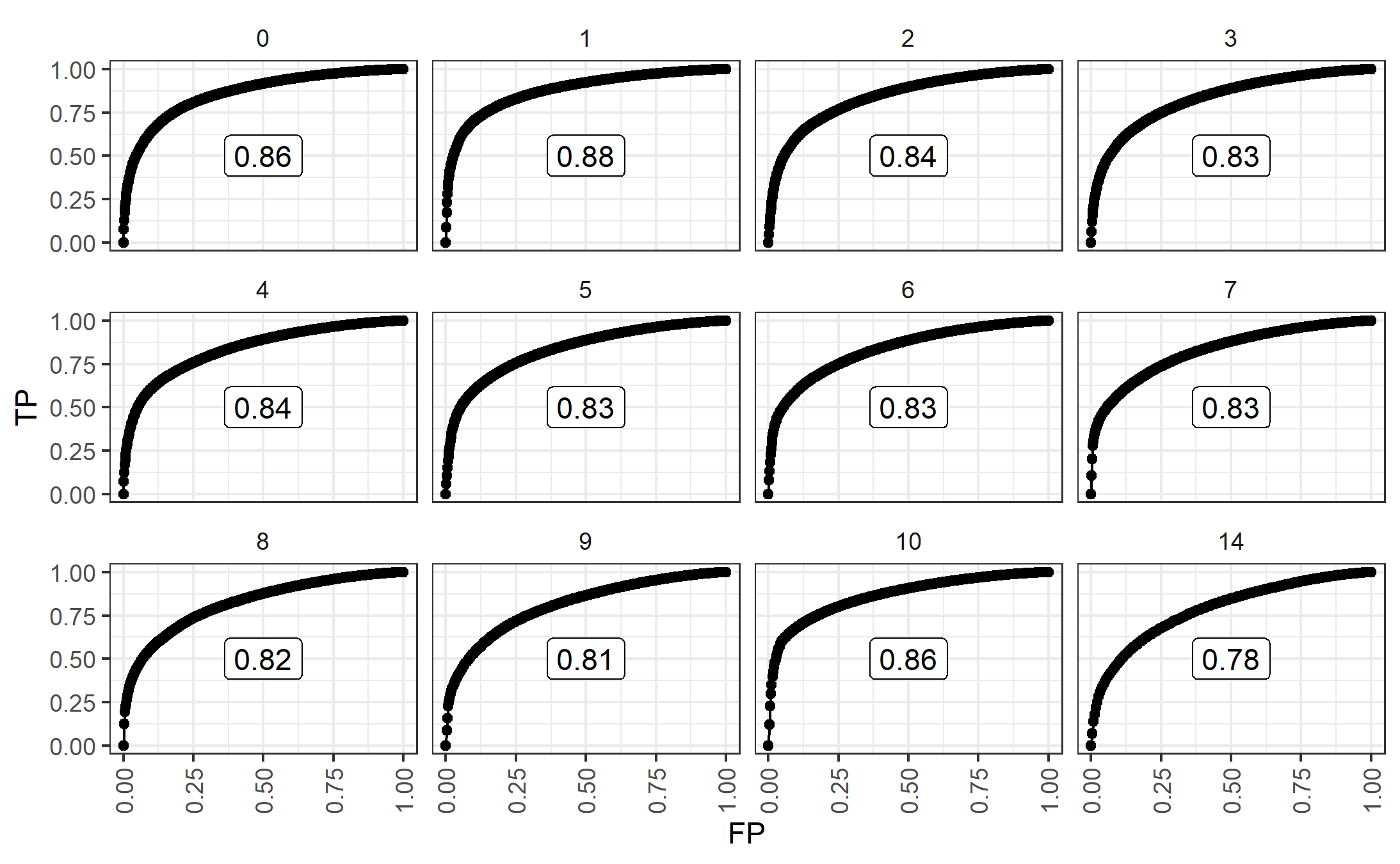

Figure S6

Baseline survival curves from fitted Cox proportional hazard models in table 3 in the derivation cohort, with 95% confidence intervals. Calculation set at mean values of cohort (model linear predictor calculations centred on mean values).

| 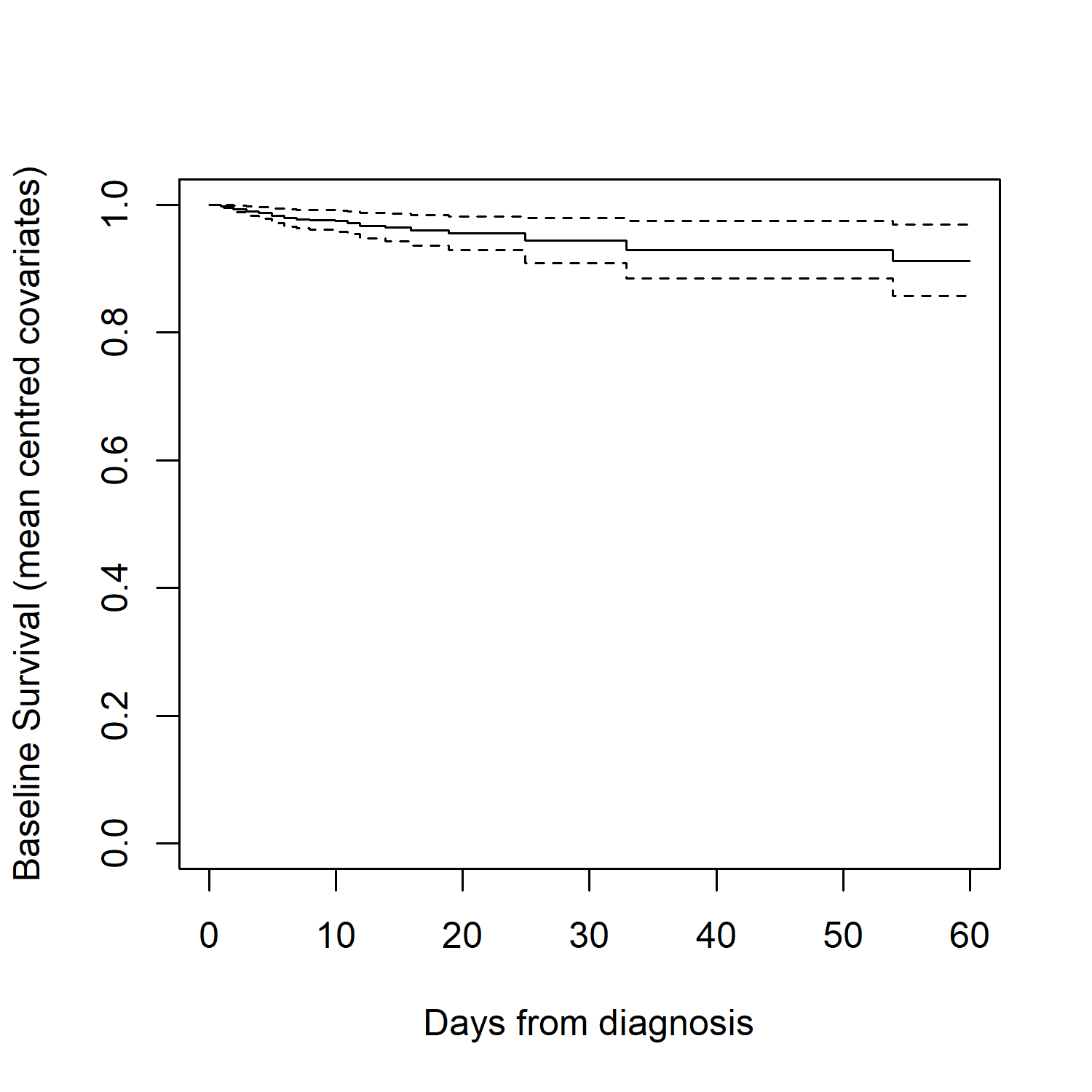 | 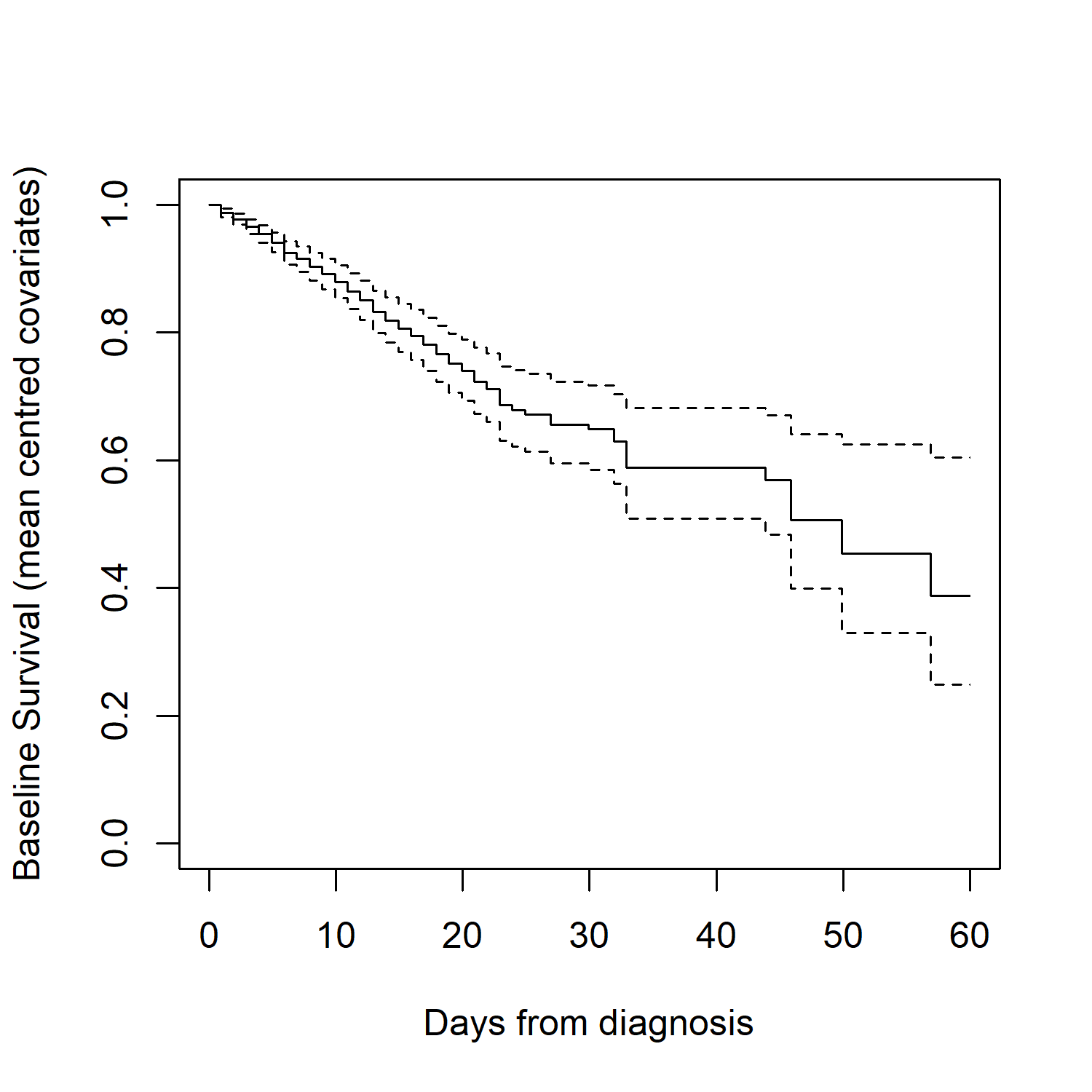 |
| --- | --- |
| (a) Patients eligible for next day escalation | (b) Patients ineligible for next day escalation |

Figure S7

Calibration curve for next day prediction of ICU admission or death in the validation cohort

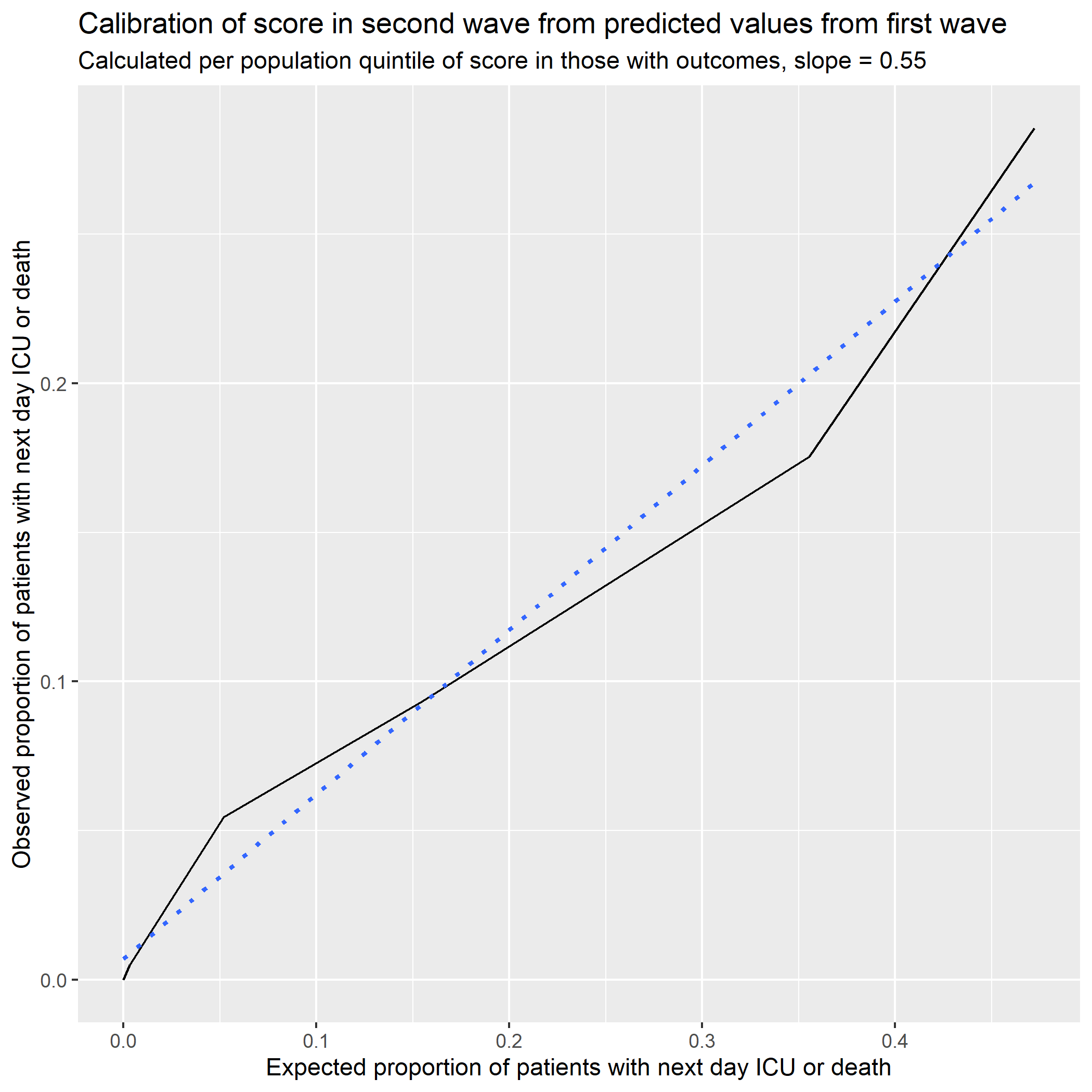

**Figure S8**

Stacked bar chart of magnitude of daily calculated linear predictor (derived in this study) overlaid with a line plot of the number of patients who were escalated or died the next day from those who were eligible for escalation (calculated using leave one out cross validation) in:

| 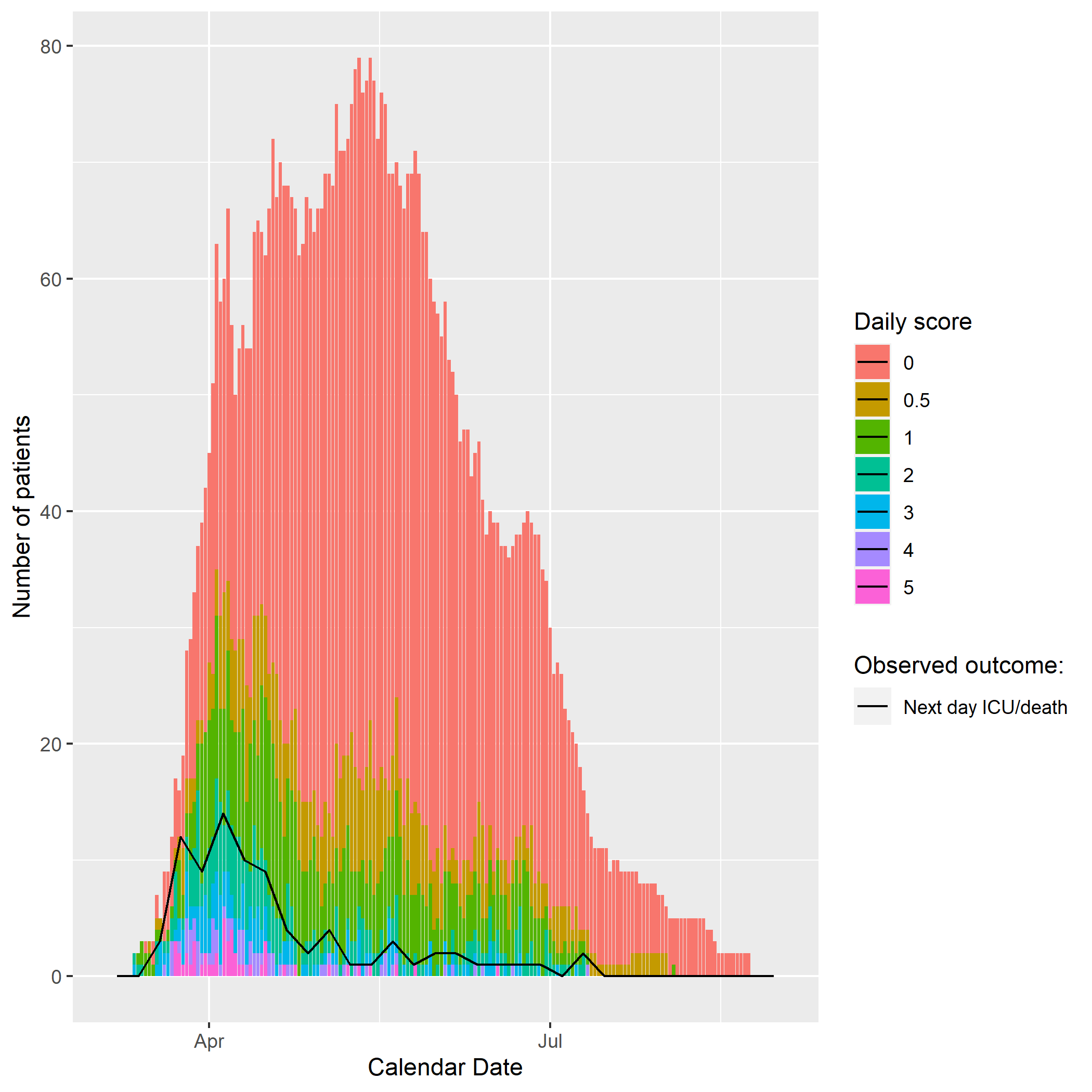 | 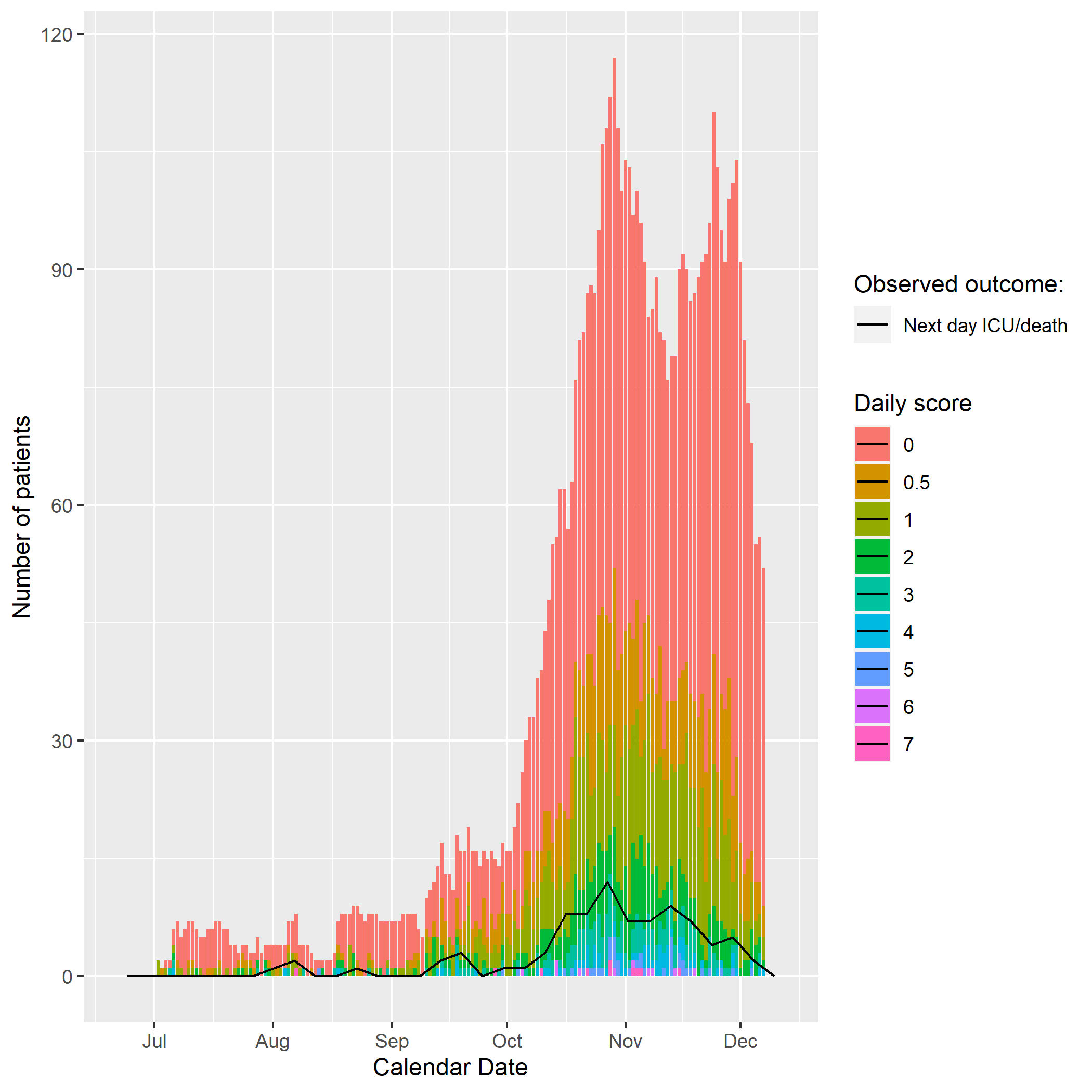 |
| --- | --- |
| (c) First wave in which score was derived   (First suspected 21 February 2020 until 30 June 2020 then followed up on a daily basis for next day events) | (d) Second wave in which score is validated   (First suspected 1^st^ July 2020 until 30 November 2020 then followed up on a daily basis for next day events) |

Figure S9

Stacked bar chart of magnitude of daily calculated linear predictor (derived in this study) overlaid with a line plot of the number of patients who died each day from those who were ineligible for escalation (calculated using leave one out cross validation) in:

| 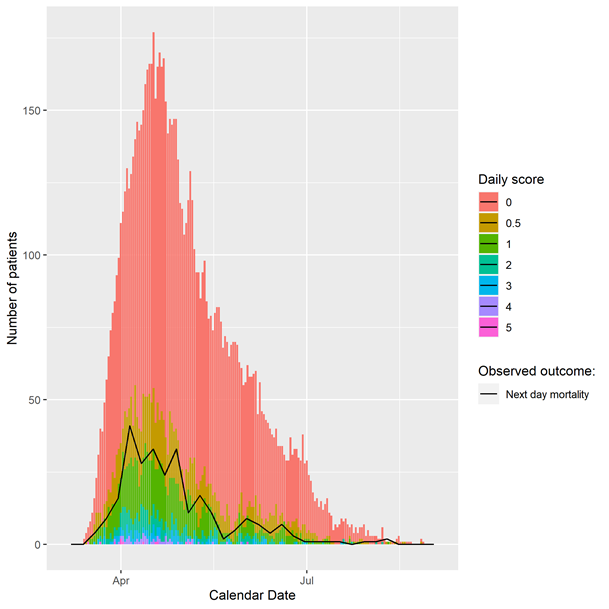 | 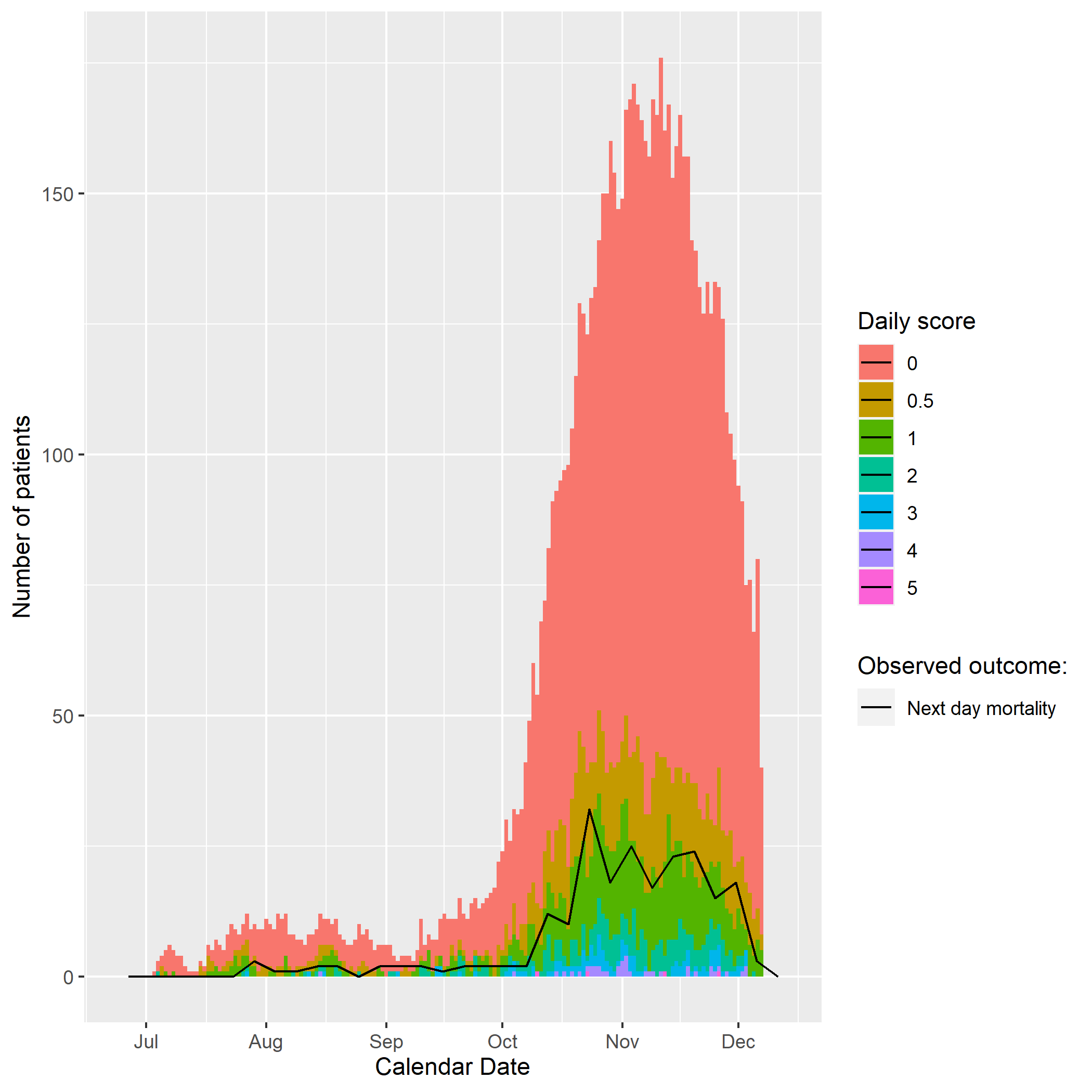 |
| --- | --- |

| (a) First wave in which score was derived   (First suspected 21 February 2020 until 30 June 2020 then followed up on a daily basis for next day events) | (b) Second wave in which score is validated   (First suspected 1^st^ July 2020 until 30 November 2020 then followed up on a daily basis for next day events) |
| --- | --- |
